# Diffusion-weighted Imaging And Retinal Oximetry Predict Functional Outcome After The First Episode Of Optic Neuritis

**DOI:** 10.1101/2025.04.03.25324536

**Authors:** Pavel Hok, Jan Valošek, Tereza Svrčinová, František Odstrčil, Martina Rybáriková, Michal Král, Kruznev S. Nijhar, Anna Arkhipova, Monika Jasenská, Jan Mareš, Martin Šín

## Abstract

**Purpose:** To evaluate diffusion weighted imaging (DWI) with advanced diffusion models, optical coherence tomography (OCT), and automatic retinal oximetry as potential biomarkers for visual deficits after optic neuritis (ON).

**Methods:** Twenty-five patients with acute unilateral ON underwent brain DWI, OCT, and oximetry at the treatment initiation, and 3 and 6 months later. Additionally, 56 healthy volunteers with normal vision were examined once. Clinical outcomes were best-corrected visual acuity (BCVA) and contrast sensitivity. Predictors included retinal nerve fiber layer (RNFL) thickness, arteriolar and venular oxygen saturation (AS, VS), and arterio-venous difference (AVD) in the affected eye as well as DWI parameters from both optic radiations.

**Results:** Multivariate analysis of variance (MANOVA) for baseline DWI indicated higher secondary partial volume fraction (f2) in patients, while both groups exhibited significant left-right differences for all DWI parameters. Longitudinal analysis in 17 patients with follow-up data revealed a multivariate effect of time when adjusted for affected side and time since onset, however, no DWI parameters changed significantly on a univariate level. Stratified unadjusted model indicated lower overall fractional anisotropy (FA) in patients with incomplete recovery. On uncorrected level, baseline FA and oximetry (VS) were correlated with follow-up BCVA at month 6, while axonal diffusivity (AD) predicted follow-up VS and AVD. In turn, baseline VS and AVD predicted follow-up RNFL thickness.

**Conclusions:** DWI and retinal oximetry are promising early predictors of visual function outcome after ON.

**Translational Relevance:** DWI and retinal oximetry are potentially suitable for patient stratification in studies investigating novel therapeutic interventions.

## 1. Introduction

Despite the general opinion that patients with optic neuritis (ON) typically achieve substantial recovery of visual function within several weeks, up to 40% of patients with ON diagnosed with multiple sclerosis (MS) exhibit abnormal best-corrected visual acuity (BCVA) or contrast sensitivity in the affected eye after 10 years.^1^ Additionally, 60% of patients report persistent difficulties after ON, potentially impacting their quality of life.^2^ Early escalation therapy with plasma exchange seems to improve outcome in case of severe ON.^3^ However, in order to identify patients profiting from new treatment strategies, early biomarkers of functional recovery after ON are required and are currently a matter of ongoing research.^2^ Retinal imaging techniques such as optical coherence tomography (OCT) have been shown to be sensitive to retinal neuronal loss and predictive of resulting visual acuity.^4^ Moreover, two OCT parameters, namely retinal nerve fiber layer (RNFL) and ganglion cell + inner plexiform layer (GCIPL), have been identified as reliable biomarkers of MS progression.^5,6^

However, retinal thinning develops gradually after acute ON due to inflammatory swelling and more immediate disease biomarkers are therefore required.^7^ Another promising retinal imaging tool – automatic retinal oximetry – has recently been used to investigate abnormalities in retinal metabolism in ON and revealed increased arteriolar oxygen saturation (AS) in the acute stage and decreased oxygen consumption 6 months after ON, as well as correlation with RNFL, potentially reflecting both acute local metabolic effects of inflammation and later neurodegeneration.^8,9^

New potential biomarkers should also reflect retrochiasmal structural abnormalities, as optic nerve inflammation cannot fully account for the decline in visual functions in patients with MS.^10^ Therefore, combination with additional diagnostic tools, such as brain diffusion weighted imaging (DWI) with diffusion tensor imaging (DTI)^11^ might provide further insight into interaction between retinal damage and white matter integrity of the optic pathway. In MS, fractional anisotropy (FA) of the optic radiation (OR) has been shown to correlate with RNFL, low contrast visual acuity^12^ or visual evoked potentials (VEP) latency.^13^ More recently, axonal diffusivity (AD) of the OR within 1 month after onset of ON was predictive for RNFL and VEP after 12 months.^14^ However, parameters obtained from more complex diffusion models,^15,16^ were shown to be more sensitive to underlying pathological processes even at the disease onset^17^ and better characterize clinical outcomes in MS.^18^ Similarly, primary and secondary partial volume fractions (f1 and f2) from the ball-and-sticks model^19^ representing measures similar to FA estimated for non-colinear axonal populations, have recently been shown to be more sensitive to tissue damage than conventional DTI parameters on the spinal level.^20,21^ It is yet unclear whether these additional DWI parameters are more sensitive to axonal damage in ON and whether they are associated with visual function outcome. Furthermore, association between DWI and retinal oximetry parameters has not yet been evaluated.

Hence, we aimed to investigate the relationship between the visual functional impairment and the integrity of the OR, as well as the interaction between the integrity of the OR and the processes occurring in the retina at different time-scales (early metabolic abnormalities in oximetry or late cell loss in OCT) in a single longitudinal study.

In detail, we hypothesized that (1) DWI parameters (FA, mean diffusivity [MD], axonal diffusivity [AD], radial diffusivity [RD], f1, f2) of the OR on each side would differ between patients with ON and healthy controls (HCs) at baseline (M0). (2) DWI parameters of the OR on each side change across follow-up (M0, month 3 [M3], and month 6 [M6]) in (a) all patients and especially in those (b) with incomplete recovery (BCVA at M6 < 20/20 or contrast sensitivity using Pelli-Robson score^22^ at M6 < 1.65 log units of the affected eye [AE]). (3) DWI parameters correlate with (a) BCVA, (b) Pelli-Robson score, (c) RNFL, and (d) retinal oximetry parameters (arteriolar saturation [AS]; venular saturation [VS], and arterio-venous difference [AVD]) of the AE at M0, M3, and M6. (4) DWI parameters at M0 predict (a) BCVA, (b) Pelli-Robson, (c) RNFL, (d) AS, VS, and AVD of the AE at M6 after ON.

Following auxiliary hypotheses were tested in patients with ON as well: (5a) RNFL, (5b) AS, VS, and AVD of the AE correlate with BCVA and Pelli-Robson AE at each time point; (6a) RNFL, (6b) AS, VS, and AVD of the AE at M0 predict ipsilateral BCVA and Pelli-Robson score at M6; (7) AS, VS, and AVD of the AE predict RNFL at M6; (8) DWI parameters correlate with lesion load (LL) at each time point.

## 2. Methods

### 2.1. Participants

Consecutive patients with the first episode of ON were recruited between 2019 and 2022 in a neurological department at a tertiary-level hospital in this longitudinal study. Inclusion criteria for patients were: age 18 to 65, history of acute unilateral ON that occurred during the previous 8 weeks with no systemic steroid treatment prior to current admission, fulfilling the criteria for clinically isolated syndrome (CIS) or relapse-remitting MS as evaluated by treating neurologist based on the 2017 revised McDonald criteria.^23^ Furthermore, two HCs were recruited per case. Inclusion criteria for HCs were: no history of any neurological or ophthalmologic condition and no signs of motor disability upon enrolment; sex and age matched to patient group. Exclusion criteria for both patients and HC were: history of diabetes, other ocular diseases, including glaucoma, refractive errors greater or equal ±6 dioptres, age-related macular degeneration, cataract, and history of vitrectomy. Additional exclusion criterion from the analysis for patients was recurrent ON. Recruitment was carried out as a part of a multimodal longitudinal imaging study and no dedicated sample size estimation was performed a priori for the here evaluated hypotheses.

The study was carried out in accordance with World Medical Association Declaration of Helsinki. Written informed consent was obtained from all participants prior to their inclusion in the study and study was approved by and conducted in compliance with the Ethics Committee of the University Hospital and the Faculty of Medicine and Dentistry, Palacký University in Olomouc, approval number NV19-06-00216.

### 2.2. Assessment Schedule

At the first visit (M0), each participant underwent standard ophthalmologic evaluation to rule out other conditions than ON, an MRI session, automatic retinal oximetry, and optical coherence tomography. Patients were scheduled for follow-up visits 3 (M3) and 6 months (M6) after the first visit repeating all assessments from M0 visit. As an effort to avoid performance bias, all patients additionally underwent standard clinical neurological assessments and received standard treatment according to national guidelines and at the discretion of the treating neurologist, including disease-modifying therapy in indicated cases. All assessments as well as definitions of variables of interest and potential confounders are provided in Supplementary Table 1.

### 2.3. Clinical Assessments

All participants were screened for history of any neurological or ophthalmologic condition. Handedness was assessed initially in all participants using Edinburgh Handedness Inventory.^24^ Ocular dominance was established at each visit using a hole-in-card procedure. The Best-Corrected Visual Acuity Measurements (BCVA) measurement was obtained using Early Treatment Diabetic Retinopathy Study (ETDRS) visual acuity chart according to the standard protocol.^25^ Contrast sensitivity was assessed using the Pelli-Robson test^22^ BCVA of <20/20 and/or the contrast sensitivity score of <1.65 log units at M6 were considered as an abnormal outcome. PwMS underwent standard neurological examination, providing disability assessment using Expanded Disability Status Scale (EDSS).^26^

### 2.4. Optical Coherence Tomography (OCT)

The retinal nerve fiber layer (RNFL) measurement was performed using high resolution spectral domain OCT (SD-OCT) combining OCT technology with a confocal scanning laser ophthalmoscope (Spectralis, software version 4.0.3.0, Heidelberg Engineering, Heidelberg, Germany) as described elsewhere.^8^ All RNFL scans were acquired after the pupil dilation allowing a detailed differentiation of retinal layers. Each eye was scanned repeatedly within one session until at least 3 high-quality RNFL scans were achieved for further analysis.

### 2.5. Automatic Retinal Oximetry

Retinal oximetry was carried out using Oxymap oximeter (Oxymap ehf., Reykjavik, Iceland). A standardized acquisition technique was employed with full description provided elsewhere.^8,27^ Image analyses was performed according to a standardized protocol (Oxymap protocol for acquisition and analysis of Oxymap T1 oximetry images, version November 21, 2013; Oxymap ehf.) using the software Oxymap Analyzer (version 3.1.4, Oxymap ehf., Reykjavik, Iceland). The arterio-venous difference (AVD) was calculated as the difference between the retinal arteriolar oxygen saturation (AS) and the retinal venular oxygen saturation (VS).^28^

### 2.6. MRI Data Acquisition

Imaging data were acquired using a 1.5 T (Siemens Aera, Erlangen, Germany) scanner with a 20-channel head/neck coil. The subject’s head was immobilized with cushions to assure comfort and minimize head motion. Imaging protocol included a high resolution three-dimensional isotropic T_1_-weighted scan using magnetization-prepared rapid acquisition with gradient echo (MPRAGE) sequence with repetition time (TR) = 2460 ms, echo time (TE) = 3.65 ms, inversion time (TI) = 1240 ms, flip angle (FA) = 8°, 224 sagittal slices with 0.5 mm gap, field of view (FOV) = 256 × 256 mm with 1 mm isotropic resolution, parallel acquisition techniques (PAT) factor = 2, fat suppression enabled; an in-plane fluid-attenuated inversion recovery (FLAIR) image with TR = 5000 ms, TE = 335 ms, TI = 1800 ms, FA = 120°, 176 sagittal slices, in-plane FOV = 224×256 mm, resolution 1 mm isotropic, PAT factor = 2; and a single-shot spin-echo echo-planar imaging (EPI) diffusion-weighted sequence (TR = 6600 ms per slice, TE = 92 ms, FA = 90°, b = 1000 s/mm^2^, bandwidth 888 Hz/Px, 64 directions covering the full sphere, 5 reference B_0_ images, FOV = 260×260 mm, 40 slices, voxel size 2 mm isotropic, and no gap between slices, PAT factor = 2, partial Fourier = 6/8, A-P phase encoding) covering the occipital and temporal lobes. A set of 6 reference B_0_ images with inverted phase encoding (i.e., P-A phase encoding) was acquired for off-line correction of B_0_ off-resonance geometrical distortions.

### 2.7. Analysis Of DWI Data

DWI data were processed using FMRIB Software Library (FSL) version 6.0.5^29^ to correct for motion, susceptibility distortions and eddy current distortions.^30,31^ Then, the conventional DTI model^11^ and multi-compartment ball-and-sticks model accounting for crossing fibers^19^ were estimated to extract microstructural parameters (FA, MD, AD, and RD for the DTI model and f1 and f2 for the ball-and-sticks model).

A two-step registration of a skull-stripped diffusion-weighted image to a standard-space MNI152 structural image^32^ via the individual structural image (MPRAGE) was carried out, providing non-linear warping field between the diffusion and MNI152 standard space. Next, the XTRACT tool^33–35^ was used to perform probabilistic tractography of the left and right OR utilizing standardized masks warped into diffusion space, hence, the tractography was performed in native space. Masks for tractography consisted of a seed mask placed in the lateral geniculate nucleus and a target mask in a coronal plane through the anterior part of the calcarine fissure. Exclusion masks consisted of an axial block of the brainstem, a coronal plane anterior to the seed, and a coronal block directly posterior to the seed.^35^ Microstructural parameters were extracted for each participant and each tract after a 5% threshold using the xtract_stats tool, effectively extracting parameters from the OR core.^35^

### 2.8. Lesion Load Assessment

Individual T_2_-weighted lesion load (LL) was calculated using the Lesion Segmentation Tool (LST, www.statistical-modelling.de/lst.html with lesion prediction algorithm.^36^ Resulting lesion maps were thresholded at *p* = 0.5. For each participant, volume fraction of the OR on each side affected by T_2_-weighted lesions was evaluated.

### 2.9. Statistical Analysis

First, normality was assessed for continuous variables using Shapiro-Wilk test. Next, potential differences in demographic (age, sex, handedness, ocular dominance), clinical (BCVA, contrast sensitivity, RNFL, retinal AS, VS, and AVD), and auxiliary imaging variables (LL) were assessed (comparison patients v. HCs and/or comparisons M0 v. M3, M0 v. M6, and M3 v. M6 where applicable). Dichotomous variables were analyzed using Fisher’s exact test for 2×2 contingency tables (Pearson Chi-Square in case of non-dichotomous nominal variables). The remaining non-normally distributed variables were compared between groups using Mann-Whitney U test, while within-group comparisons were carried out using Wilcoxon signed rank test. Main hypotheses 1 and 2 were tested using doubly multivariate analysis of variance (MANOVA) with DWI parameters (FA, MD, AD, RD, f1, f2) for the left and right OR as dependent variables and the hemisphere (side of the OR) as a within-subject factor. After confirming that dependent variables did not deviate significantly from normal distribution, bivariate Pearson correlation for each pair of variables was calculated to assess potential multicollinearity (|*r*| > 0.9). This was the case for correlations between FA and f1, RD and f1, and partly between FA and RD. For the sake of comparability with previous studies reporting FA, the two remaining offending variables (f1 and RD) were discarded from the main analyses. However, to evaluuate how the results were affected by this arbitrary choice, we conducted a sensitivity analysis, in which the MANOVA was additionally repeated after excluding FA and RD while including f1.

For hypothesis 1, patient and HC data at M0 were compared with group as a between-subject factor. To assess the robustness of results, two additional models were evaluated: first, sex and age were added as covariates; second, a patients were compared to one-to-one matched HCs selected using SPSS matching algorithm for sex, age, ocular dominance, and handedness. For hypothesis 2a, patient data for M0, M3, and M6 were compared with each other with time as a within-subject factor. For hypothesis 2b, abnormal outcome at M6 (BCVA < 20/20 and/or Pelli-Robson score < 1.65 log of the AE) was added as a between-subject factor. In an additional model, robustness of results was assessed by adding time since onset at M0 and the affected side (left or right) as additional covariates.

Each MANOVA was interpreted according to the following procedure^37^: First, multivariate Wilk’s lambda test was examined. For each significant multivariate test (*p* < 0.05), corresponding post-hoc univariate statistics (i.e., *F* tests resulting from a series of one-way analyses of variance [ANOVA]) were assessed for each dependent variable (Bonferroni-corrected *p* < 0.0125). In case of a significant multivariate interaction with a significant corresponding post-hoc univariate interaction, simple mean effects were evaluated. In case of significant multivariate main effects without interaction, pair-wise comparisons with Sidak correction were carried out for factor time, or for dichotomous factors, corresponding univariate *F* tests were reported as final outcomes (Bonferroni-corrected *p* < 0.0125).

For the remaining hypotheses, bivariate correlations were assessed within the patient group using Spearman rank correlation coefficient. First, relationship between DWI parameters (FA, MD, AD, RD, f1, f2) and ophthalmological assessments (BCVA, Pelli-Robson score, RNFL, AS, VS, AVD) was evaluated at corresponding time points (hypothesis 3). Next, correlation between the baseline DWI parameters (M0) and the follow-up ophthalmology assessments (M6) was calculated (hypothesis 4). For auxiliary hypotheses 5-7, association between visual function (BCVA and Pelli-Robson of the affected side) and ipsilateral retinal parameters (RNFL, AS, VS, and AVD) was evaluated at corresponding time points (hypotheses 5), as well as between baseline retinal parameters and follow-up visual function (hypothesis 6) or RNFL (hypothesis 7). Finally, correlation between DWI parameters and LL including lesion fraction of the OR was assessed at corresponding time points (hypothesis 8). Correlations for main hypotheses were considered significant at *p* < 0.05 with additional Bonferroni-Holm correction for number of independent variables.

All analyses were performed in SPSS Statistics 29 (IBM, Armonk, NY, USA, https://www.ibm.com/analytics/spss-statistics-software) with alpha significance level *p* < 0.05. Missing data were excluded pairwise.

### 2.10. Additional Post-hoc Analyses

Due to low variability of BCVA and contrast sensitivity score at M6, an additional receiver operating characteristic (ROC) analysis was carried out for prediction of aggregate abnormal outcome at M6 (BCVA < 20/20 and/or Pelli-Robson score < 1.65 log of the AE) with the DWI parameters (FA) and ophthalmological assessments (VS) significantly associated with any of the outcome measures used as predictors.^38^ Uncorrected asymptotic significance at *p* < 0.05 was considered significant for null hypothesis area under curve (AUC) = 0.5, with paired-sample AUC difference considered significant at *p* < 0.05 under null hypothesis AUD difference = 0.

## 3. Results

### 3.1. Study Sample

In total, 33 consecutive patients with ON and 68 HCs were enrolled. Subsequently, 12 HCs were excluded due to incidental abnormal ophthalmological findings (3), abnormal BVCA (4), missing ophthalmological data (3), large susceptibility artifact (1), and corrupt imaging data preventing further analysis (1). One patient was excluded due to history of diabetes and 7 patients were excluded from the analysis due to missing MRI data for following reasons: claustrophobia (4), refusal to undergo study MRI without providing the reason (2), gastrointestinal infection at baseline resulting in withdrawal (1).

Out of the remaining 25 patients, 1 patient had missing baseline DWI data, but completed ophthalmological assessments and was included in correlation analysis for M3 and M6. Furthermore, 1 patient withdrew prior to M3 visit (no reason provided) and 3 patients had an ON relapse within follow-up period and have thus been completely excluded from the longitudinal analysis (but were included in baseline analysis). Furthermore, 1 patient missed M6 MRI (but completed ophthalmological assessments and was included in correlation analysis for M0, M3, and prediction of M6 outcome) and 2 patients withdrew due to pregnancy after M3 and prior to M6 (but were included in the baseline analysis and correlation analysis for M3). Demographic parameters and group comparisons of included subjects are provided in Table 1. Overall, there were no significant differences between the patient and HC groups. Individual clinical characteristics are provided in Table 2.

**Table 1.** Demographic characteristics and group comparisons.

|  | Patients<br>(*n* = 25) | HC (matched)<br>(*n* = 25) | *P* | HC<br>(*n* = 56) | *P* |
| --- | --- | --- | --- | --- | --- |
| Age [years]* | 32.8 ±14.3 | 30.3 ±9.6 | 0.691<sup>†</sup> | 29.2 ±5.8 | 0.481<sup>†</sup> |
| Sex (women/men) | 20/5 | 20/5 | 1.000<sup>‡</sup> | 37/19 | 0.293<sup>‡</sup> |
| Ocular dominance<br>(L/R/missing) | 8/17/0 | 8/17/0 | 1.000<sup>§</sup> | 13/40/3 | 0.536<sup>§</sup> |
| Handedness LI* | 0.9 ±0.3 | 0.8 ±0.3 | 0.889<sup>†</sup> | 0.8 ±0.3 | 0.765<sup>†</sup> |
| Affected side (L/R) | 6/19 | N/A | N/A | N/A | N/A |

**Notes:** \*)Median ±interquartile range; <sup>†</sup>)Mann-Whitney U-test; <sup>‡</sup>)Fisher's exact test; <sup>§</sup>)Pearson Chi-Square (exact).
**Abbreviations:** HC – healthy controls; L – left; LI – (Edinburgh Handedness Inventory) laterality index; N/A – not applicable; R – right.

**Table 2.** Clinical patient characteristics.

| *n* | Age range | Sex | Affected eye | Time onset-MRI M0 [d] | Time corticoids-MRI M0 [d]* | BCVA (decimal) |  |  | Pelli-Robson [log units] |  |  | Diagnosis | Relapses |  | DMT | Dropout |
| --- | --- | --- | --- | --- | --- | --- | --- | --- | --- | --- | --- | --- | --- | --- | --- | --- |
|  |  |  |  |  |  | M0 | M3 | M6 | M0 | M3 | M6 | M0 | *n* | ON | M6 |  |
| 1 | 21-25 | w | R | 7 | 1 | 1.00 | 1.00 | N/A | N/A | N/A | N/A | CIS | 0 |  | none | WD M6 – pregnancy |
| 2 | 26-30 | w | R | 28 | -1 | 0.80 | 0.80 | 1.00 | N/A | N/A | N/A | CIS | 0 |  | none |  |
| 3 | ≤ 20 | w | L | 4 | 0 | 0.16 | N/A | 1.00 | N/A | N/A | N/A | CIS | 0 |  | none |  |
| 4 | 21-25 | w | R | 7 | -1 | 0.03 | 0.80 | 0.63 | N/A | N/A | 1.65 | CIS | 1 | no | none |  |
| 5 | 36-40 | w | R | 54 | -1 | 0.32 | 0.40 | 0.25 | N/A | N/A | N/A | CIS | 0 |  | none |  |
| 6 | 31-35 | m | L | 15 | -1 | 0.40 | N/A | 1.00 | N/A | N/A | 1.95 | CIS | 1 | yes | none |  |
| 7 | 21-25 | w | R | 4 | -2 | N/A | 0.63 | N/A | N/A | 1.80 | N/A | CIS | 0 |  | IFN-β | WD M6 – pregnancy |
| 8 | 21-25 | w | L | 6 | 0 | 1.00 | 1.00 | 1.00 | N/A | N/A | 1.95 | CIS | 0 |  | IFN-β |  |
| 9 | 41-45 | w | L | 13 | 7 | 0.20 | 0.50 | 0.50 | N/A | 1.35 | 1.35 | CIS | 1 | yes | N/A |  |
| 10 | 36-40 | w | L | 32 | 1 | 0.63 | 0.00 | 0.00 | 1.20 | 0.30 | 0.00 | CIS | 1 | yes | none |  |
| 11 | 41-45 | w | L | 9 | 0 | 0.70 | N/A | N/A | 0.60 | N/A | N/A | CIS | N/A |  | none | WD M3 – no reason |
| 12 | 36-40 | w | R | 21 | 5 | 1.00 | 1.00 | 1.00 | 1.65 | 1.80 | 1.95 | CIS | 0 |  | TFL |  |
| 13 | 21-25 | w | R | 7 | 2 | 0.80 | 0.80 | 1.00 | 1.50 | 1.35 | 1.35 | CIS | 0 |  | IFN-β |  |
| 14 | 31-35 | m | L | 13 | 3 | 0.80 | 1.00 | 1.00 | 1.50 | 1.80 | 1.80 | CIS | 0 |  | IFN-β |  |
| 15 | 31-35 | m | L | 13 | 1 | 1.00 | 1.00 | 1.00 | 1.95 | 1.95 | 1.95 | CIS | 0 |  | TFL |  |
| 16 | ≤ 20 | w | L | 11 | 0 | 0.03 | 0.80 | 1.00 | 0.60 | 1.95 | 1.95 | CIS | 0 |  | IFN-β |  |
| 17 | 26-30 | w | L | 11 | 7 | 1.00 | 1.00 | 1.00 | 1.95 | 1.95 | 1.95 | CIS | 0 |  | TFL |  |
| 18 | 26-30 | w | L | 22 | -3 | 1.00 | 1.00 | 1.00 | 1.50 | 1.95 | 1.95 | CIS | 0 |  | none |  |
| 19 | 36-40 | w | L | 7 | 4 | 0.12 | 1.00 | 1.00 | 0.60 | 1.60 | 1.65 | CIS | 0 |  | TFL |  |
| 20 | ≤ 20 | m | L | 10 | 3 | 0.32 | 1.00 | 0.80 | 0.75 | 1.65 | 1.95 | CIS | 0 |  | OFA |  |
| 21 | 46-50 | w | L | 7 | 3 | 0.05 | 1.00 | 1.00 | 0.00 | 1.80 | 1.95 | CIS | 0 |  | IFN-β |  |
| 22 | 31-35 | m | L | 2 | 0 | 0.80 | 1.00 | 1.00 | 1.05 | 1.95 | 1.95 | CIS | 0 |  | IFN-β |  |
| 23 | 26-30 | w | L | 11 | 2 | 0.63 | 1.00 | 1.00 | 1.35 | 1.80 | 1.95 | CIS | 2 | no | OFA |  |
| 24 | 41-45 | w | R | 54 | 17 | 1.00 | 1.00 | 1.00 | 1.65 | 1.65 | 1.65 | CIS | 0 |  | OFA |  |
| 25 | 36-40 | w | R | 11 | 1 | 0.03 | 0.05 | 0.05 | 0.15 | 0.30 | 0.45 | CIS | 0 |  | TFL |  |

**Notes:** \*)positive values indicate corticoid treatment initiated before magnetic resonance imaging;
**Abbreviations:** CIS – clinically isolated syndrome; DMT – disease modifying therapy; IFN-β – interferon β1a; L – left; m – man; M0 – month 0 (baseline); M3 – month 3; M6 – month 6; N/A – not applicable; OFA – ofatumumab; ON – optic neuritis; R – right; rON – recurrent optic neuritis; RRMS – relapsing-remitting multiple sclerosis; SD – standard deviation; w – woman; TFL – teriflunomide; yrs – years; WD – withdrawn.

### 3.2. Clinical Assessment

Both BCVA and contrast sensitivity in the AE were significantly decreased in patients at baseline (median decimal BCVA = 0.67; median Pelli-Robson score = 1.28 log units) compared to HC with normal vision (Table 3). While contrast sensitivity significantly improved at each time point (median Pelli-Robson score was 1.80 and 1.95 log units for M3 and M6, respectively), a significant improvement in BCVA was only observed between M0 and M3 (median BCVA was 1.00 for both M3 and M6), see Table 3 for statistical comparisons. In total, 5 patients (i.e., 26.3% out of 19 patients included in the follow-up analysis) still exhibited abnormal vision of the AE at M6, i.e., had BCVA < 20/20 or Pelli-Robson score < 1.65 log.

**Table 3.** Visual function and retinal imaging, group comparisons.

|  |  | Patients – affected eye |  |  |  |  | HC – random eye |  |  |  |
| --- | --- | --- | --- | --- | --- | --- | --- | --- | --- | --- |
|  |  | Median | IQR | *n* | *p* v. M3*<br>(*n*) | *p* v. M6*<br>(*n*) | Median | IQR | *n* | *p* v. PwON† |
| Decimal BCVA | M0‡ | 0.67 | 0.83 | 24 | 0.005 (19) | 0.002 (19) | 1.00 | 0.00 | 56 | <0.001 |
|  | M3§ | 1.00 | 0.20 | 20 |  | 0.518 (18) |  |  |  | <0.001 |
|  | M6§ | 1.00 | 0.00 | 19 |  |  |  |  |  | 0.001 |
| Pelli-Robson score | M0‡ | 1.28 | 1.01 | 16 | 0.006 (14) | 0.004 (14) | 1.95 | 0.00 | 39 | <0.001 |
|  | M3§ | 1.80 | 0.30 | 15 |  | 0.024 (14) |  |  |  | <0.001 |
|  | M6§ | 1.95 | 0.30 | 16 |  |  |  |  |  | 0.092 |
| RNFL [µm] | M0‡ | 101.5 | 19 | 24 | 0.003 (19) | 0.003 (19) | N/A | N/A | 0 | N/A |
|  | M3§ | 96.5 | 24 | 20 |  | 0.047 (18) |  |  |  | N/A |
|  | M6§ | 87.0 | 23 | 19 |  |  |  |  |  | N/A |
| Retinal AS [%] | M0‡ | 98 | 4 | 23 | 0.931 (17) | 0.391 (18) | 97 | 3 | 56 | 0.125 |
|  | M3§ | 98 | 3 | 19 |  | 0.806 (17) |  |  |  | 0.309 |
|  | M6§ | 98 | 7 | 19 |  |  |  |  |  | 0.063 |
| Retinal VS [%] | M0‡ | 66 | 8 | 23 | 0.434 (17) | 0.571 (18) | 65 | 7 | 56 | 0.378 |
|  | M3§ | 65 | 9 | 19 |  | 0.962 (17) |  |  |  | 0.898 |
|  | M6§ | 67 | 10 | 19 |  |  |  |  |  | 0.893 |
| Retinal AVD [%] | M0‡ | 31 | 10 | 23 | 0.619 (17) | 0.631 (18) | 32 | 6 | 56 | 0.725 |
|  | M3§ | 32 | 8 | 19 |  | 0.669 (17) |  |  |  | 0.669 |
|  | M6§ | 33 | 8 | 19 |  |  |  |  |  | 0.709 |

Notes: \*)Wilcoxon Signed Rank Test; †)Mann-Whitney U-test; ‡)all included subjects; §)PwON included in follow-up analyses (*n* = 21).
**Abbreviations:** AS – arteriolar (oxygen) saturation; AVD – arterio-venous difference; BCVA – Best-Corrected Visual Acuity; IQR – interquartile range; L – left; N/A – not applicable; PwON – patients with optic neuritis; R – right; RNFL – retinal nerve fiber layer; VS – venular (oxygen) saturation.

We observed significant gradual decline in median RNFL thickness in the AE, decreasing from 101.5 μm at M0 to 87.0 μm at M6 (Table 3). In contrast, we found no difference in retinal oxygen saturation levels between the patients and HCs (affected eye vs. random eye) at baseline and observed no change in the AE in patients during follow-up, see Table 3.

### 3.3. Lesion Load Assessment

Median ±IQR of T2-weighted LL for all included patients at baseline (n = 25) was 175 ±632 mm^3^. For patients included in follow-up analysis, it was 192 ±760 mm^3^ at M0, 156 ±471 mm^3^ at M3, and 248 ±645 mm^3^ at M6. No paired-sample differences in LL were detected (uncorrected *p_M0-M3_*= 0.167, *p_M0-M6_*= 0.278, *p_M3-M6_*= 0.868, Wilcoxon Signed Rank Test). Individual lesion volume fraction in the OR ranged from 0% to 4.0% (median = IQR = 0% for both left and right OR at each time point), hence whole fiber tracts were assessed in the statistical analysis.

### 3.4. Differences In DWI At Baseline

In total 24 patients were eligible for MANOVA for group differences in DWI parameters at baseline (hypothesis 1). Summary statistics are provided in Supplementary Table 2. In the unadjusted model, MANOVA yielded significant multivariate effects of group (i.e., difference between patients with ON and HCs) and the hemisphere while no significant interaction was observed. For the factor hemisphere, post-hoc ANOVA was significant for each tested dependent variable (FA, MD, AD, and f2) showing that higher FA, MD, AD and lower f2 were detected in the left OR compared to the right OR independent of group membership. For the group factor, univariate ANOVA yielded significant effect for f2 (higher f2 in patients compared to HCs), but not for the remaining dependent variables, see Table 4 and Fig. 1.

**Fig. 1.**
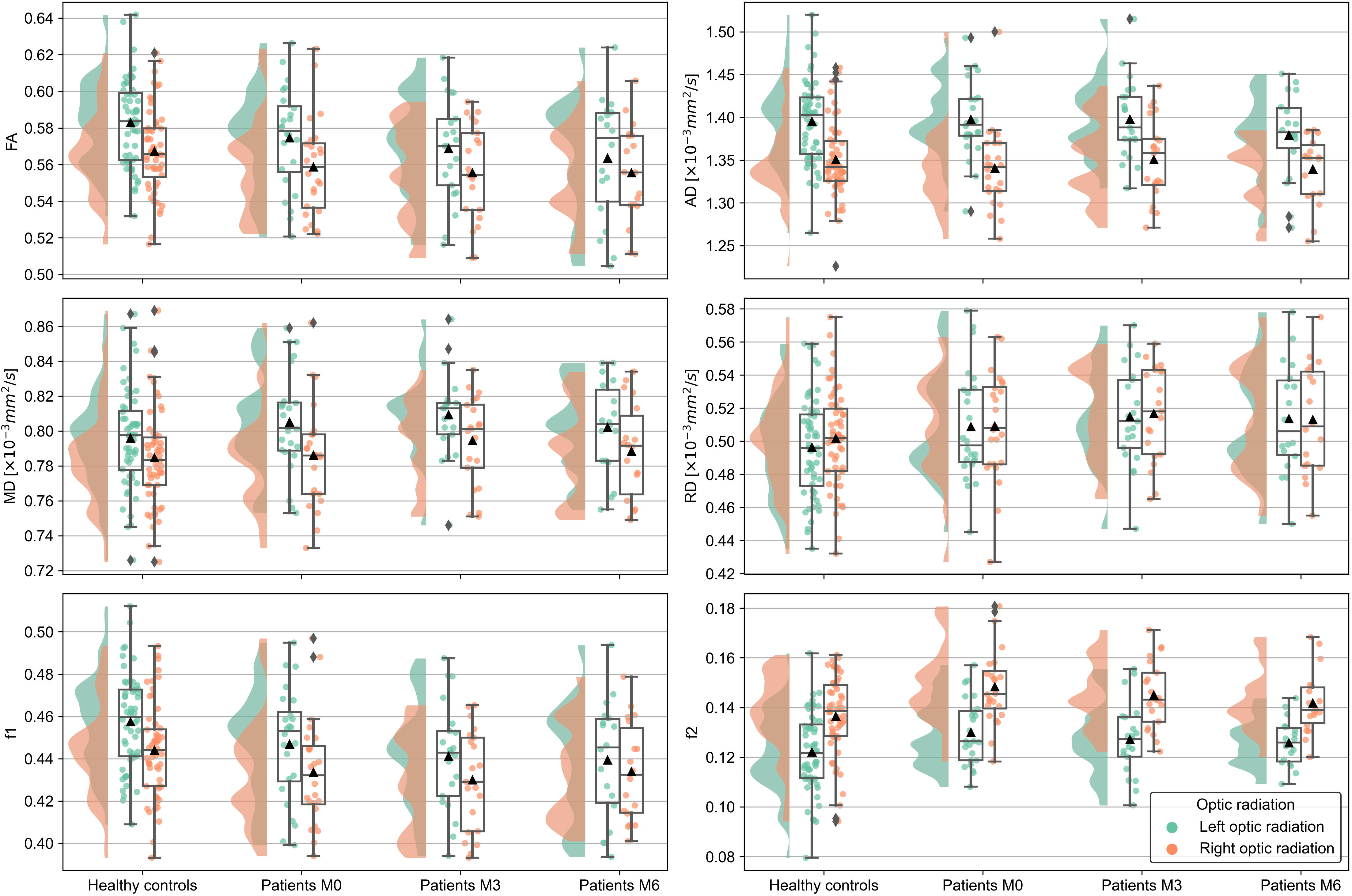
Diffusion-weighted imaging (DWI) parameters by group and visit. Raincloud plots in all panels show distribution as well as individual values for fractional anisotropy (FA), axial, mean and radial diffusivity (AD, MD, RD, respectively), primary and secondary partial volume fractions (f1 and f2, respectively) in healthy controls and patients with optic neuritis at month 0, 3, and 6 (M0, M3, and M6, respectively). Data from the left (green – left) and right (red – right) optic radiation are displayed separately. The embedded box plots additionally provide median, quartiles and outliers, whereas mean is indicated by a triangle.

**Table 4.** MANOVA and post-hoc ANOVA for group comparison at baseline.

| Model | factor | MANOVA |  |  |  |  | Variable | ANOVA |  |  |  |
| --- | --- | --- | --- | --- | --- | --- | --- | --- | --- | --- | --- |
| | | Wilk's $\lambda$ | df* | $F$ | $p$ | Partial $\eta^2$ | | df* | $F$ | $p^\dagger$ | Partial $\eta^2$ |
| Unadjusted<br>24 patients vs. 56 HCs | group | 0.842 | 4, 75 | 3.526 | 0.011 | 0.158 | FA | 1, 78 | 2.503 | 0.118 | 0.031 |
|  |  |  |  |  |  |  | MD | 1, 78 | 0.639 | 0.427 | 0.008 |
|  |  |  |  |  |  |  | AD | 1, 78 | 0.148 | 0.702 | 0.002 |
|  |  |  |  |  |  |  | f2 | 1, 78 | 8.799 | **0.004** | 0.101 |
|  | hemisphere | 0.310 | 4, 75 | 41.729 | <0.001 | 0.690 | FA | 1, 78 | 34.210 | <0.001 | 0.305 |
|  |  |  |  |  |  |  | MD | 1, 78 | 49.453 | <0.001 | 0.388 |
|  |  |  |  |  |  |  | AD | 1, 78 | 138.484 | <0.001 | 0.640 |
|  |  |  |  |  |  |  | f2 | 1, 78 | 81.230 | <0.001 | 0.510 |
|  | group*hemisphere | 0.930 | 4, 75 | 1.415 | 0.237 | 0.070 |  |  |  |  |  |
| Adjusted<br>24 patients vs. 56 HCs | group | 0.905 | 4, 72 | 1.894 | 0.121 | 0.095 |  |  |  |  |  |
|  | hemisphere | 0.937 | 4, 72 | 1.220 | 0.310 | 0.063 |  |  |  |  |  |
|  | age | 0.935 | 4, 72 | 1.260 | 0.294 | 0.065 |  |  |  |  |  |
|  | sex | 4.798 | 4, 72 | 0.790 | **0.002** | 0.210 | FA | 1, 75 | 2.752 | 0.101 | 0.035 |
|  |  |  |  |  |  |  | MD | 1, 75 | 0.415 | 0.521 | 0.006 |
|  |  |  |  |  |  |  | AD | 1, 75 | 4.185 | 0.044 | 0.053 |
|  |  |  |  |  |  |  | f2 | 1, 75 | 16.643 | <0.001 | 0.182 |
|  | group*sex | 0.968 | 4, 72 | 0.599 | 0.665 | 0.032 |  |  |  |  |  |
|  | group*hemisphere | 0.951 | 4, 72 | 0.921 | 0.457 | 0.049 |  |  |  |  |  |
|  | hemisphere*age | 0.991 | 4, 72 | 0.163 | 0.956 | 0.009 |  |  |  |  |  |
|  | hemisphere*sex | 0.994 | 4, 72 | 0.117 | 0.976 | 0.006 |  |  |  |  |  |
|  | group*hemisphere*sex | 0.991 | 4, 72 | 0.169 | 0.953 | 0.009 |  |  |  |  |  |
| Unadjusted<br>24 patients vs. 24 HCs | group | 0.771 | 4, 43 | 3.199 | **0.022** | 0.229 | FA | 1, 46 | 4.780 | 0.034 | 0.094 |
|  |  |  |  |  |  |  | MD | 1, 46 | 0.533 | 0.469 | 0.011 |
|  |  |  |  |  |  |  | AD | 1, 46 | 1.266 | 0.266 | 0.027 |
|  |  |  |  |  |  |  | f2 | 1, 46 | 7.795 | **0.008** | 0.145 |
|  | hemisphere | 0.303 | 4, 43 | 24.704 | <0.001 | 0.697 | FA | 1, 46 | 28.181 | <0.001 | 0.380 |
|  |  |  |  |  |  |  | MD | 1, 46 | 41.767 | <0.001 | 0.476 |
|  |  |  |  |  |  |  | AD | 1, 46 | 93.242 | <0.001 | 0.670 |
|  |  |  |  |  |  |  | f2 | 1, 46 | 48.213 | <0.001 | 0.512 |
|  | group*hemisphere | 0.872 | 4, 43 | 1.572 | 0.199 | 0.128 |  |  |  |  |  |

Notes: \*)Order: hypothesis df, error df; <sup>†</sup>)Bonferroni-corrected significance level $p < 0.0125$ for $n = 4$ marked in **bold**.
**Abbreviations:** AD – axonal diffusivity; ANOVA – analysis of variance; df – degrees of freedom; f2 – secondary partial volume fraction; FA – fractional anisotropy; HCs – healthy controls; MANOVA – multivariate ANOVA; MD – mean diffusivity.

The analysis with matched HC confirmed significant multivariate effects of group and hemisphere. For the factor group, significant univariate effect for f2 was detected (FA was significant at uncorrected level), with higher f2 observed in patients compared to HCs, see Table 4. However, in the adjusted model with sex and age as covariates, neither effect for factor group nor effect for factor hemisphere remained significant on multivariate level. Instead, a significant multivariate effect of sex was obtained, with post-hoc univariate test indicating significant effects for f2 on the corrected significance level, with lower f2 in men compared to women, see Table 4 and Fig. 2. Finally, the results of the corresponding analyses including f1 (and excluding FA) mirrored the results of the main analyses with FA, see Supplementary Table 3.

**Fig. 2.**
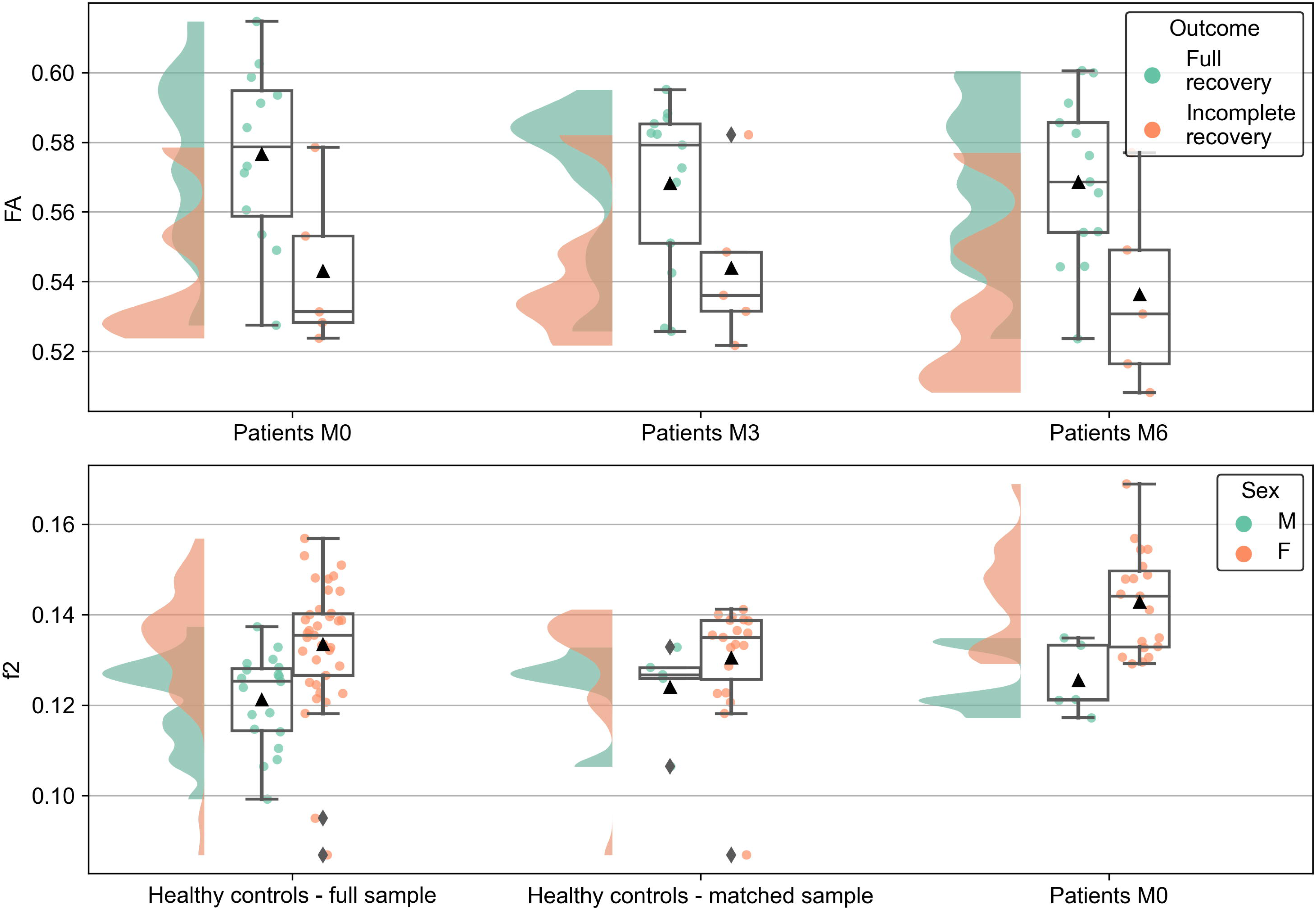
Diffusion-weighted imaging (DWI) parameters grouped by outcome and sex. Raincloud plots show distribution as well as individual values for fractional anisotropy (FA, **top panel**) and for secondary partial volume fraction (f2, **bottom panel**). In both panels, values for the right and left hemisphere were averaged. In the **top panel**, patient data for month 0, 3, and 6 (M0, M3, and M6, respectively) are sorted according to complete (green – left) or incomplete (red – right) visual function recovery, defined as best-corrected visual acuity (BCVA) < 20/20 and/or Pelli-Robson score < 1.65 log of the affected eye at M6. In the **bottom panel**, data in healthy controls (both full sample with n = 56 and matched sample with n = 24) and patients at M0 were grouped by sex (green – left for men [M] and red – right for women [F]). As in Fig. 1, the embedded box plots show median, quartiles and outliers, whereas mean is indicated by a triangle.

### 3.5. Longitudinal Changes In DWI

In total, 17 cases were eligible for MANOVA to assess changes in DWI parameters in patients during follow-up (hypothesis 2a). In the unadjusted model, MANOVA yielded significant multivariate effects for the hemisphere (i.e., difference between the left and right OR) and interaction time*hemisphere. For this interaction, Mauchly’s test of sphericity indicated sphericity assumption violation for FA (*p* = 0.041), hence, Greenhouse-Geisser correction with *ε* = 0.743 was applied. The post-hoc ANOVA for time*hemisphere with Greenhouse-Geisser correction for FA yielded no significant effect for any of the variables (Table 5), hence main effect of the hemisphere was examined, yielding significant post-hoc F tests for MD, AD, f2, but not for FA (Table 5 and Fig. 1).

**Table 5.** MANOVA and post-hoc ANOVA for longitudinal assessment in patients, irrespective of outcome.

| Model | factor | MANOVA |  |  |  |  | ANOVA |  |  |  |  |
| --- | --- | --- | --- | --- | --- | --- | --- | --- | --- | --- | --- |
| | | Wilk's $\lambda$ | df* | $F$ | $p$ | Partial $\eta^2$ | Variable | df* | $F$ | $p^\dagger$ | Partial $\eta^2$ |
| Unadjusted | time | 0.580 | 8, 9 | 0.815 | 0.608 | 0.420 |  |  |  |  |  |
|  | hemisphere | 0.219 | 4, 13 | 11.599 | <0.001 | 0.781 | FA | 1, 16 | 5.151 | 0.037 | 0.244 |
|  |  |  |  |  |  |  | MD | 1, 16 | 15.308 | **0.001** | 0.489 |
|  |  |  |  |  |  |  | AD | 1, 16 | 38.641 | **<0.001** | 0.707 |
|  |  |  |  |  |  |  | f2 | 1, 16 | 38.017 | **<0.001** | 0.704 |
|  | time*hemisphere | 0.245 | 8, 9 | 3.473 | **0.041** | 0.755 | FA<sup>‡</sup> | 1.5, 23.8 | 0.680 | 0.474 | 0.041 |
|  |  |  |  |  |  |  | MD | 2, 32 | 1.549 | 0.228 | 0.088 |
|  |  |  |  |  |  |  | AD | 2, 32 | 1.040 | 0.365 | 0.061 |
|  |  |  |  |  |  |  | f2 | 2, 32 | 0.582 | 0.565 | 0.035 |
| Adjusted | time | 0.163 | 8, 7 | 4.485 | **0.031** | 0.837 | FA | 2, 28 | 3.708 | 0.037 | 0.209 |
|  |  |  |  |  |  |  | MD | 2, 28 | 2.555 | 0.096 | 0.154 |
|  |  |  |  |  |  |  | AD | 2, 28 | 4.294 | 0.024 | 0.235 |
|  |  |  |  |  |  |  | f2 | 2, 28 | 0.580 | 0.566 | 0.040 |
|  | hemisphere | 0.426 | 4, 11 | 3.709 | **0.038** | 0.574 | FA | 1, 14 | 0.058 | 0.813 | 0.004 |
|  |  |  |  |  |  |  | MD | 1, 14 | 13.039 | **0.003** | 0.482 |
|  |  |  |  |  |  |  | AD | 1, 14 | 12.035 | **0.004** | 0.462 |
|  |  |  |  |  |  |  | f2 | 1, 14 | 6.136 | 0.027 | 0.305 |
|  | affected side | 0.603 | 4, 11 | 1.814 | 0.196 | 0.397 |  |  |  |  |  |
|  | time since onset | 0.825 | 4, 11 | 0.585 | 0.680 | 0.175 |  |  |  |  |  |
|  | time*hemisphere | 0.269 | 8, 7 | 2.380 | 0.135 | 0.731 |  |  |  |  |  |
|  | time*affected side | 0.387 | 8, 7 | 1.389 | 0.339 | 0.613 |  |  |  |  |  |
|  | time*<br>time since onset | 0.177 | 8, 7 | 4.057 | **0.041** | 0.823 | FA | 2, 28 | 2.396 | 0.109 | 0.146 |
|  |  |  |  |  |  |  | MD | 2, 28 | 1.945 | 0.162 | 0.122 |
|  |  |  |  |  |  |  | AD | 2, 28 | 2.886 | 0.073 | 0.171 |
|  |  |  |  |  |  |  | f2 | 2, 28 | 0.694 | 0.508 | 0.047 |
|  | hemisphere*<br>affected side | 0.864 | 4, 11 | 0.431 | 0.783 | 0.136 |  |  |  |  |  |
|  | hemisphere*<br>time since onset | 0.703 | 4, 11 | 1.161 | 0.379 | 0.297 |  |  |  |  |  |
|  | time*hemisphere*<br>affected side | 0.200 | 8, 7 | 3.502 | 0.058 | 0.800 |  |  |  |  |  |
|  | time*hemisphere*<br>time since onset | 0.316 | 8, 7 | 1.894 | 0.207 | 0.684 |  |  |  |  |  |

**Notes:** \*)Order: hypothesis df, error df; <sup>†</sup>)Bonferroni-corrected significance level $p < 0.0125$ for $n = 4$ marked in **bold**; <sup>‡</sup>)Greenhouse-Geisser corrected univariate statistics.
**Abbreviations:** AD – axonal diffusivity; ANOVA – analysis of variance; df – degrees of freedom; f2 – secondary partial volume fraction; FA – fractional anisotropy; MANOVA – multivariate ANOVA; MD – mean diffusivity.

In the remaining models, no sphericity violation was detected (*p* > 0.05, Mouchly’s test of sphericity). For the adjusted model for changes in DWI parameters with time since onset and affected side as covariates, MANOVA yielded significant multivariate effects of time, hemisphere and interaction time*time since onset. The post-hoc ANOVA for time*time since onset yielded no significant effect for any of the variables (Table 5), hence main effect of time and hemisphere were examined, yielding significant post-hoc *F* tests for the factor hemisphere in case of MD and AD (Table 5 and Fig. 1). For the factor time, no significant F tests were detected after Bonferroni correction (on uncorrected level, FA and AD were significant).

In the model for changes in DWI parameters with stratification according to outcome (but no further covariates) for hypothesis 2b, MANOVA yielded significant multivariate effects of abnormal outcome, hemisphere and interaction time*hemisphere. The post-hoc ANOVA for time*hemisphere yielded no significant effect for any of the variables (Table 6), hence main effects of abnormal outcome and hemisphere were examined, yielding significant post-hoc F tests for the factor hemisphere in case of MD, AD, and f2 (Table 6). For the factor abnormal outcome, one significant *F* test was detected after Bonferroni correction for FA, with lower FA in patients with abnormal outcome (Fig. 2).

**Table 6.** MANOVA and ANOVA for longitudinal assessment in patients, stratified by outcome.

| Model | factor | MANOVA |  |  |  |  | ANOVA |  |  |  |  |
| --- | --- | --- | --- | --- | --- | --- | --- | --- | --- | --- | --- |
| | | Wilk's $\lambda$ | df* | $F$ | $p$ | Partial $\eta^2$ | Variable | df* | $F$ | $p^\dagger$ | Partial $\eta^2$ |
| Stratified by outcome | abnormal outcome | 0.304 | 4, 12 | 6.883 | **0.004** | 0.696 | FA | 1, 15 | 8.058 | **0.012** | 0.349 |
|  |  |  |  |  |  |  | MD | 1, 15 | 0.424 | 0.525 | 0.027 |
|  |  |  |  |  |  |  | AD | 1, 15 | 3.037 | 0.102 | 0.168 |
|  |  |  |  |  |  |  | f2 | 1, 15 | 1.566 | 0.230 | 0.095 |
|  | time | 0.641 | 8, 8 | 0.560 | 0.785 | 0.359 |  |  |  |  |  |
|  | hemisphere | 0.263 | 4, 12 | 8.396 | **0.002** | 0.737 | FA | 1, 15 | 3.911 | 0.067 | 0.207 |
|  |  |  |  |  |  |  | MD | 1, 15 | 11.436 | **0.004** | 0.433 |
|  |  |  |  |  |  |  | AD | 1, 15 | 28.808 | **<0.001** | 0.658 |
|  |  |  |  |  |  |  | f2 | 1, 15 | 27.090 | **<0.001** | 0.644 |
|  | abnormal outcome*<br>time | 0.636 | 8, 8 | 0.571 | 0.777 | 0.364 |  |  |  |  |  |
|  | abnormal outcome*<br>hemisphere | 0.856 | 4, 12 | 0.503 | 0.734 | 0.144 |  |  |  |  |  |
|  | time*hemisphere | 0.153 | 8, 8 | 5.529 | **0.013** | 0.847 | FA | 2, 30 | 1.763 | 0.189 | 0.105 |
|  |  |  |  |  |  |  | MD | 2, 30 | 1.041 | 0.365 | 0.065 |
|  |  |  |  |  |  |  | AD | 2, 30 | 1.270 | 0.295 | 0.078 |
|  |  |  |  |  |  |  | f2 | 2, 30 | 0.855 | 0.435 | 0.054 |
|  | abnormal outcome*<br>time*hemisphere | 0.253 | 8, 8 | 2.947 | 0.074 | 0.747 |  |  |  |  |  |

Notes: \*)Order: hypothesis df, error df; <sup>†</sup>Bonferroni-corrected significance level $p < 0.0125$ for $n = 4$ marked in **bold**.
Abbreviations: AD – axonal diffusivity; ANOVA – analysis of variance; df – degrees of freedom; f2 – secondary partial volume fraction; FA – fractional anisotropy; MANOVA – multivariate ANOVA; MD – mean diffusivity.

The results of the longitudinal analysis including FA were largely reproduced in the additional analysis using corresponding models in which FA was replaced by f1 (Supplementary Table 4 and 5). However, several differences were observed: First, the unadjusted model yielded no significant multivariate interaction time*hemisphere. Second, the model adjusted for affected side and time since onset yielded no significant interaction time*time since onset. Instead, a multivariate interaction time*affected side was detected, but it was not significant on the univariate level (Supplementary Table 4). Finally, the model stratified by outcome yielded no significant univariate effect of abnormal outcome (although f1 was significant on uncorrected level; Supplementary Table 5).

### 3.6. Correlations

Spearman rank correlation coefficient for relationship between DWI parameters and ophthalmological assessments (BCVA, Pelli-Robson score, RNFL, AS, VS, AVD, i.e., hypothesis 3) yielded no significant correlation after Bonferroni-Holm-correction at *p* < 0.0083. For full results, see Supplementary Table 6.

The correlation between the baseline DWI parameters (M0) and the follow-up ophthalmology assessments at M6 (hypothesis 4) yielded significant correlation between AD in the left OR and follow-up VS of the AE (Spearman’s *ρ* = −0.616, *p* = 0.006, *n* = 18) at Bonferroni-Holm-correction at *p* < 0.0083, while on uncorrected level, higher baseline FA tended to be associated with higher BCVA and Pelli-Robson at M6, see Table 7.

**Table 7.**
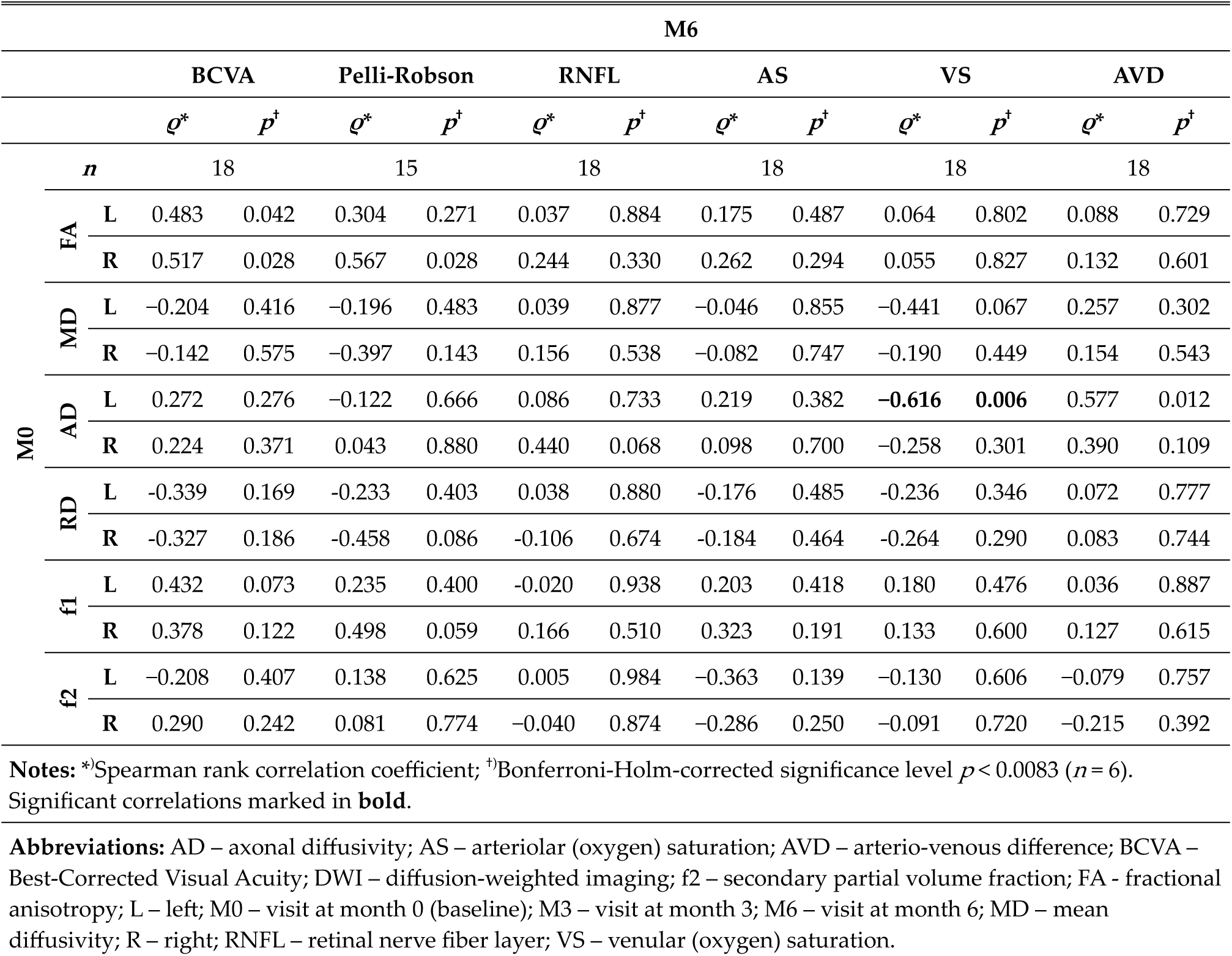
Prediction of ophthalmological parameters at M6 by baseline parameters.

For auxiliary hypotheses 5-7, uncorrected two-tailed Spearman rank correlation yielded significant negative correlation between AVD and BCVA at baseline (*ρ* = −0.604, *p* = 0.003, *n* = 22), and positive correlation between RNFL and BCVA at M6 (*ρ* = 0.495, *p* = 0.031, *n* = 19). Retinal VS of the AE significantly predicted BCVA at M6 (*ρ* = 0.609, *p* = 0.007, *n* = 18) and similar trend was observed for AVD (*ρ* = −0.410, *p* = 0.091, *n* = 18). Both VS and AVD of the AE significantly predicted RNFL at M6 (VS: *ρ* = 0.571, *p* = 0.013; AVD: *ρ* = −0.747, *p* = <0.001, *n* = 18), see Supplementary Table 7 for full results. Finally, no significant correlation between DWI parameters and LL (including lesion volume fraction of the OR) was observed at any time point at *p* < 0.05 uncorrected (hypothesis 8), see Supplementary Table 8.

### 3.7. Additional Analyses

The post-hoc ROC analysis was performed for FA of the left and right OR, and for VS of the AE. We observed above-chance AUC for all three tested predictors (FA of the right OR: AUC = 0.850, *p* = 0.002; FA of the left OR: AUC = 0.767, *p* = 0.043; and VS of the AE: AUC = 0.792, *p* = 0.023, *n* = 17, asymptotic two-tailed significance for null hypothesis AUC = 0.5), see Fig. 3. No significant paired-sample differences were observed among the three ROC curves.

**Fig. 3.**
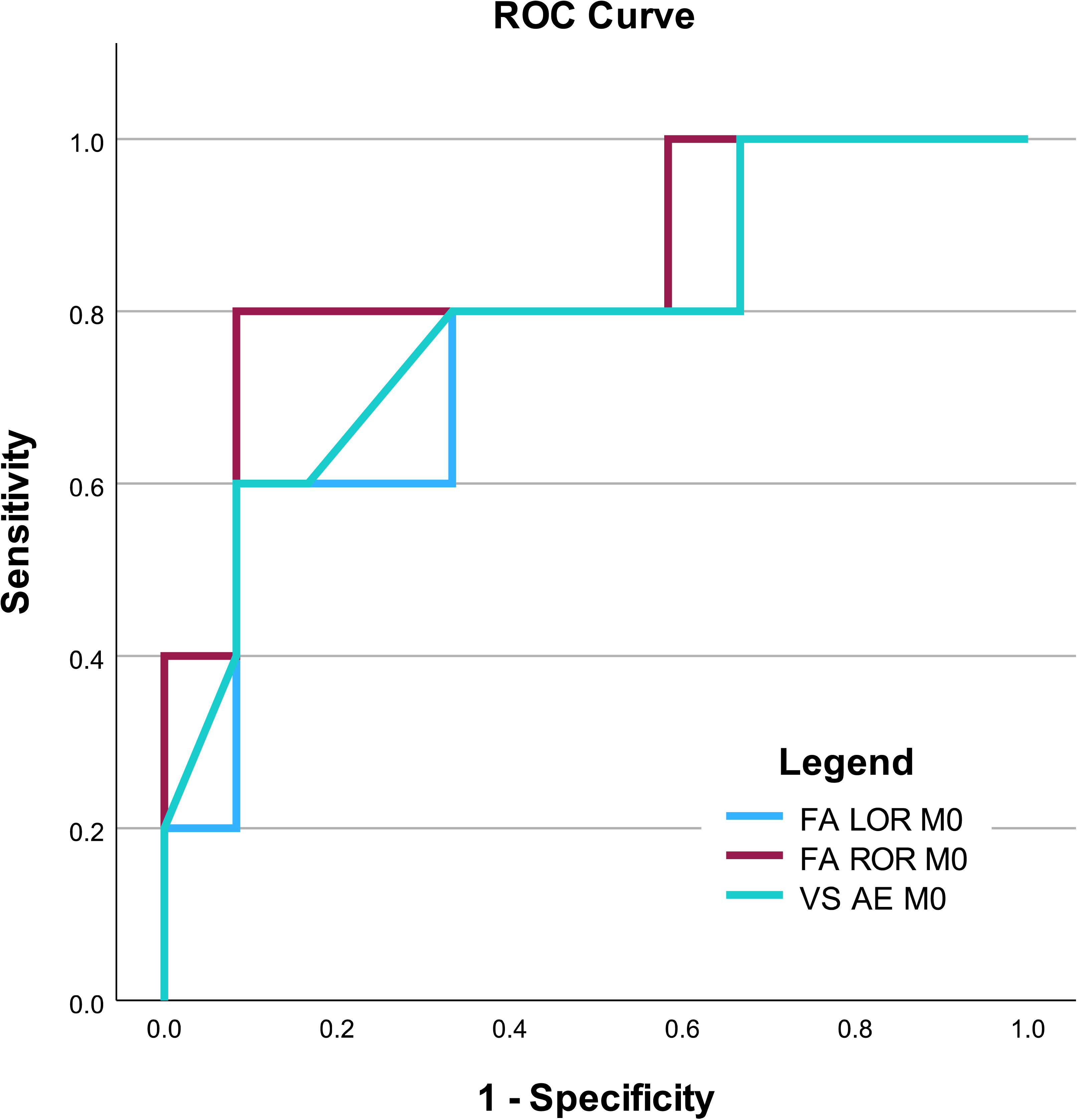
Receiver operating characteristic (ROC) curve for prediction of incomplete recovery. The ROC curve illustrates sensitivity vs. 1 − specificity for fractional anisotropy (FA) of the left and right optic radiation (LOR and ROR, respectively) and retinal venular saturation (VS) of the affected eye (AE) at baseline (M0) predicting abnormal outcome at month 6, i.e., the best-corrected visual acuity (BCVA) < 20/20 and/or Pelli-Robson score < 1.65 log of the AE.

## 4. Discussion

### 4.1. Baseline Group Differences In DWI Parameters

While we observed significantly higher f2 in patients compared to controls in the sex-matched healthy control subgroup, no parameters derived from the standard DTI model (FA, MD, AD) differed between the patient and HC groups (FA, however, differed on uncorrected level in the matched sample). Similarly, previous studies found no or only a minimal difference in FA in the OR in patients with acute or early ON.^39–42^ This indicates that abnormalities in classical DTI parameters of the OR are not prominent features of acute ON, although decreases in FA have been reported in normal appearing white matter (NAWM) of the OR in patients 1-4 years after ON,^43–46^ or in MS regardless of ON history.^12,47,48^

To our knowledge, there has been no prior study assessing DWI parameters in the OR after ON using ball-and-sticks diffusion model. In this study, differences in f1 between groups were only observed on uncorrected level, i.e., similar to FA. In fact, FA and f1 were strongly correlated with each other, suggesting that f1 conveys little to no additional information on top of FA. Interestingly, we observed group differences in f2, which represents the secondary fiber population within each voxel, i.e., crossing, kissing, or fanning fibers.^20,49^ We speculate that increased proportion of crossing fibers in patients with ON could result from a relative loss of the primary fiber population oriented parallel to the OR, which could precede the decrease of overall FA observed later in the disease course.^44,46^ The finding of sex-related differences in f2 in the adjusted model suggests, however, pre-existing microstructural sexual dimorphism in the OR similar to previously reported sex-related differences in classical DTI parameters.^50^ Moreover, prominent lateralization was detected here for all DWI parameters across both groups. Although a systematic methodological bias (e.g., due to asymmetric tractography) cannot be ruled out, corresponding left-right differences in the OR have been reported before.^50,51^ Taken together, the observed lateralization and sexual dimorphism hinder the interpretability of the group differences in f2 and indicate that its use as a disease biomarker would be challenging.

### 4.2. Longitudinal Changes In DWI Parameters

Several previous studies showed gradual decline in FA the OR following ON, but no changes for AD or MD.^40,42^ Our data confirmed significant multivariate change in DWI parameters over time, with post-hoc *F* tests indicating possible effects for FA and AD and a trend for MD on uncorrected level. However, this was only observed when adjusted for the time of ON onset at M0 visit (with significant multivariate interaction time*time since onset), suggesting possible non-linear decline of DWI parameters. Although Tur et al.^42^ observed no evidence of non-linear effects, future studies should consider non-linear trajectory and/or time since symptom onset as a covariate when assessing DWI parameters after ON.

Importantly, our model stratified according to outcome identified significantly lower FA in patients who had incomplete recovery after 6 months, but indicated no interaction abnormal outcome*time. This indicates that patients who are still affected 6 months after ON have overall lower FA, but their rate of FA decline does not differ substantially from those with a complete recovery. However, a statistical trend for interaction abnormal outcome*time*hemisphere (Table 6) was observed. Therefore, a possible significant interaction in a larger sample cannot be ruled out.

### 4.3. Correlation Of DWI With Visual Function And Retinal Atrophy

Contrary to our expectations, neither visual function outcomes nor RNFL were significantly correlated with DWI at the respective time points. However, the lack of significant correlation is generally in line with previous literature in comparable cohorts. Several studies in patients after ON found no significant correlations of DWI parameters in the OR with visual acuity or RNFL.^39,41,46–48^ In another study, change in RNFL correlated with annualized change in AD, but not in FA.^40^ However, studies in larger samples including patients with MS regardless of ON history found significant correlations of FA with RNFL^12,45,52^ and visual acuity when controlling for DWI parameters outside the OR.^12^ Hence, larger sample sizes and/or longer follow-up seem to be warranted to confirm relationship between DWI parameters, visual acuity and RNFL thinning after ON.

As in case of cross-sectional correlations, we observed no significant relationship between baseline DWI and follow-up visual function assessment or RNFL at the corrected significance level. However, due to mutual correlation of multiple independent variables tested here, application of Bonferroni correction might be too conservative. Here, a biologically plausible relationship can be inferred based on a consistent pattern rather than a statistical significance.^12^ In fact, we detected several such consistent associations between the visual function outcome (both BCVA and contrast sensitivity) and baseline FA at uncorrected level. Because of a low number of abnormal findings in BCVA and contrast sensitivity assessments at M6, performance of the predictors was evaluated in an additional ROC analysis using a dichotomized aggregate visual function outcome. Here, FA provided a good discrimination (AUC of 0.850 and 0.767 for the right and left OR, respectively) between patients with complete and incomplete recovery. To our knowledge, no DWI-based predictors of visual outcome after acute ON have so far been identified in the OR.^42^ Although a recent study has revealed that AD and RD at 1 or 3 months after ON onset were predictive for RNFL at the subsequent 6-month and 12-month follow-up visits, no prediction of visual acuity at 6-month or 12-month visit was detected.^14^ Hence, independent validation of FA as a predictor of visual outcome in a larger dataset is still required.

### 4.4. Correlation Between DWI Parameters And Retinal Oximetry

Despite the lack of significant cross-sectional correlations, we detected a significant negative association between baseline AD in the left OR and follow-up retinal VS in the affected eye (corrected for number of predicted variables). Our previous study in a larger cohort of patients with acute ON revealed a decrease in AVD along with a trend for increased VS in the affected and fellow eyes after a 6-month follow-up, which was suggested to reflect the retinal atrophy.^8^ The potential mechanism linking DWI and oximetry thus might involve neurodegeneration as decreases in AD have also been associated with axonal loss.^53^ The histopathological interpretation of AD remains, however, challenging when mixed pathology is expected as in the case of MS,^53^ rendering the interpretation to a large degree speculative. The consistent correlations at M0 and M6 indicate, nevertheless, that the early microstructural features of the OR that are predictive for follow-up AVD are preserved throughout the 6-month follow-up period.

### 4.5. Correlations Among Retinal Oximetry, Visual Function, And RNFL

Our auxiliary analyses yielded several interrelated findings. The baseline AVD was correlated with baseline visual impairment. Baseline AVD and VS also predicted the follow-up RNFL, while VS also predicted follow-up visual acuity (and a trend was detected for AVD). Finally, the BCVA and RNFL at M6 were also correlated with each other. Taken together, these correlations suggest that increased oxygen metabolism at baseline (as reflected by lower VS and higher AVD)^8,27^ corresponds to the extent of inflammation and the resulting acute and residual vision impairment and retinal atrophy. These are novel results as the predictive utility of retinal oximetry for functional outcome or RNFL thinning after ON has not been assessed before. The AUC for VS was comparable with AUC for FA, suggesting that both baseline parameters are promising potential biomarkers of visual function recovery. Future studies should assess the combined performance of FA and VS (here, logistic regression analysis was not feasible due to insufficient sample size).

### 4.6. Limitations And Possible Sources Of Bias

Several limitations have to be acknowledged. First, a relatively small study sample could have negatively affected the power of our tests. The sample size was affected by a large drop out rate due to refusal to undergo MRI, which was possibly related to ongoing COVID-19 pandemics. Nevertheless, previous studies in similar cohorts with acute neuritis had comparable sample sizes^42^ and our significant results indicate sufficient sample size for large-sized effects. Furthermore, in this study the whole OR in each participant was assessed. While we did not observe any correlation with LL, local effects in the OR due to intersecting T2 lesions cannot be ruled out and a future assessment of NAWM is warranted. As in each DWI study, selection of the fiber tract of interest is challenging. Here, robust and reproducible method using widely available toolbox (XTRACT) was employed in order to maximize replicability of our results. Lastly, we have deliberately ignored the effect of treatment. In our cohort, each patient received systemic corticoids at M0, hence, our results cannot be generalized to untreated subjects. Disease modifying treatment was initiated in some patients at the discretion of treating physician as per local guidelines. Due to heterogeneity of initiated treatments, an analysis of treated vs. untreated patients was not deemed informative. Observational studies in larger cohorts and possibly interventional studies are warranted to establish the effect of early treatment initiation in ON and to test whether results can be generalized to any disease modifying treatment.

## 5. Conclusions

While optic neuritis (ON) was not associated with microstructural changes in the optic radiations (OR) that would consistently affect all patients, incomplete recovery of the visual functions at the six-month follow-up was associated with lower baseline fractional anisotropy (FA) and increased retinal oxygen metabolism. Retinal oximetry additionally predicted subsequent development of retinal nerve fiber layer (RNFL) atrophy. Our results suggest possible utility of DWI and oximetry for stratification of patients with ON in future studies investigating novel therapeutic interventions.

## Data Availability

Anonymized preprocessed data are available via a request to the corresponding author, following a formal data sharing agreement.

## Acknowledgments

The authors thank Mr. Martin Slíva for statistical consultation and Dr. Dalibor Zimek for assistance with patient recruitment. The authors also thank Prof. Jan Kremláček and Dr. Matthias Grothe for their valuable comments on an earlier version of the manuscript.

## References

1. Beck RW, Gal RL, Bhatti MT, et al. Visual function more than 10 years after optic neuritis: experience of the optic neuritis treatment trial. Am J Ophthalmol. 2004;137(1):77–83.

2. Jenkins TM, Toosy AT. Optic neuritis: the eye as a window to the brain. Curr Opin Neurol. 2017;30(1):61–66. doi:10.1097/WCO.0000000000000414

3. Galetta K, Ryan S, Manzano G, et al. Treatment outcomes of first-ever episode of severe optic neuritis. Mult Scler Relat Disord. 2022;66:104020. doi:10.1016/j.msard.2022.104020

4. Chan JW. Recent advances in optic neuritis related to multiple sclerosis. Acta Ophthalmol. 2012;90(3):203–209. doi:10.1111/j.1755-3768.2011.02145.x

5. Montolío A, Cegoñino J, Garcia-Martin E, Pérez Del Palomar A. Comparison of Machine Learning Methods Using Spectralis OCT for Diagnosis and Disability Progression Prognosis in Multiple Sclerosis. Ann Biomed Eng. 2022;50(5):507–528. doi:10.1007/s10439-022-02930-3

6. Petzold A, Balcer LJ, Calabresi PA, et al. Retinal layer segmentation in multiple sclerosis: a systematic review and meta-analysis. Lancet Neurol. 2017;16(10):797–812. doi:10.1016/S1474-4422(17)30278-8

7. Galetta SL, Villoslada P, Levin N, et al. Acute optic neuritis. Neurology Neuroimmunology & Neuroinflammation. 2015;2(4):e135. doi:10.1212/NXI.0000000000000135

8. Svrčinová T, Hok P, Šínová I, et al. Changes in oxygen saturation and the retinal nerve fibre layer in patients with optic neuritis associated with multiple sclerosis in a 6-month follow-up. Acta Ophthalmologica. 2020;98(8):841–847. 10.1111/aos.14463

9. Svrčinová T, Mareš J, Chrapek O, et al. Changes in oxygen saturation and the retinal nerve fibre layer in patients with optic neuritis - a pilot study. Acta Ophthalmol. 2018;96(3):e309–e314. doi:10.1111/aos.13571

10. Alshowaeir D, Yannikas C, Garrick R, et al. Multifocal VEP assessment of optic neuritis evolution. Clinical Neurophysiology. 2015;126(8):1617–1623. doi:10.1016/j.clinph.2014.11.010

11. Basser PJ, Mattiello J, LeBihan D. MR diffusion tensor spectroscopy and imaging. Biophys J. 1994;66(1):259–267. doi:10.1016/S0006-3495(94)80775-1

12. Reich DS, Smith SA, Gordon-Lipkin EM, et al. Damage to the optic radiation in multiple sclerosis is associated with retinal injury and visual disability. Arch Neurol. 2009;66(8):998–1006. doi:10.1001/archneurol.2009.107

13. Lobsien D, Ettrich B, Sotiriou K, Classen J, Then Bergh F, Hoffmann KT. Whole-brain diffusion tensor imaging in correlation to visual-evoked potentials in multiple sclerosis: a tract-based spatial statistics analysis. AJNR Am J Neuroradiol. 2014;35(11):2076–2081. doi:10.3174/ajnr.A4034

14. Nguyen MNL, Zhu C, Kolbe SC, et al. Early predictors of visual and axonal outcomes after acute optic neuritis. Front Neurol. 2022;13:945034. doi:10.3389/fneur.2022.945034

15. Jeurissen B, Tournier JD, Dhollander T, Connelly A, Sijbers J. Multi-tissue constrained spherical deconvolution for improved analysis of multi-shell diffusion MRI data. NeuroImage. 2014;103:411–426. doi:10.1016/j.neuroimage.2014.07.061

16. Zhang H, Schneider T, Wheeler-Kingshott CA, Alexander DC. NODDI: practical in vivo neurite orientation dispersion and density imaging of the human brain. Neuroimage. 2012;61(4):1000–1016. doi:10.1016/j.neuroimage.2012.03.072

17. Collorone S, Prados F, Kanber B, et al. Brain microstructural and metabolic alterations detected in vivo at onset of the first demyelinating event. Brain. 2021;144(5):1409–1421. doi:10.1093/brain/awab043

18. Storelli L, Pagani E, Meani A, Preziosa P, Filippi M, Rocca MA. Advanced diffusion-weighted imaging models better characterize white matter neurodegeneration and clinical outcomes in multiple sclerosis. J Neurol. 2022;269(9):4729–4741. doi:10.1007/s00415-022-11104-z

19. Behrens TEJ, Woolrich MW, Jenkinson M, et al. Characterization and propagation of uncertainty in diffusion-weighted MR imaging. Magn Reson Med. 2003;50(5):1077–1088. doi:10.1002/mrm.10609

20. Labounek R, Valošek J, Horák T, et al. HARDI-ZOOMit protocol improves specificity to microstructural changes in presymptomatic myelopathy. Scientific Reports. 2020;10(1):undefined-undefined. doi:10.1038/s41598-020-70297-3

21. Valošek J, Labounek R, Horák T, et al. Diffusion magnetic resonance imaging reveals tract-specific microstructural correlates of electrophysiological impairments in non-myelopathic and myelopathic spinal cord compression. Eur J Neurol. 2021;28(11):3784–3797. doi:10.1111/ene.15027

22. Pelli DG, Robson JG, Wilkins AJJ. The design of a new letter chart for measuring contrast sensitivity. Clinical Vision Sciences. Published online 1988:187–199.

23. Thompson AJ, Banwell BL, Barkhof F, et al. Diagnosis of multiple sclerosis: 2017 revisions of the McDonald criteria. The Lancet Neurology. 2018;17(2):162–173. doi:10.1016/S1474-4422(17)30470-2

24. Oldfield RC. The assessment and analysis of handedness: the Edinburgh inventory. Neuropsychologia. 1971;9(1):97–113.

25. Ferris FL, Kassoff A, Bresnick GH, Bailey I. New visual acuity charts for clinical research. Am J Ophthalmol. 1982;94(1):91–96.

26. Kurtzke JF. Rating neurologic impairment in multiple sclerosis: an expanded disability status scale (EDSS). Neurology. 1983;33(11):1444–1452. doi:10.1212/wnl.33.11.1444

27. Král M, Svrčinová T, Hok P, et al. Correlation between retinal oxygen saturation and the haemodynamic parameters of the ophthalmic artery in healthy subjects. Acta Ophthalmologica. 2022;100(7):e1489–e1495. doi:10.1111/aos.15189

28. Traustason S, Jensen AS, Arvidsson HS, Munch IC, Søndergaard L, Larsen M. Retinal oxygen saturation in patients with systemic hypoxemia. Invest Ophthalmol Vis Sci. 2011;52(8):5064–5067. doi:10.1167/iovs.11-7275

29. Jenkinson M, Beckmann CF, Behrens TEJ, Woolrich MW, Smith SM. FSL. Neuroimage. 2012;62(2):782–790. doi:10.1016/j.neuroimage.2011.09.015

30. Andersson JLR, Skare S, Ashburner J. How to correct susceptibility distortions in spin-echo echo-planar images: application to diffusion tensor imaging. Neuroimage. 2003;20(2):870–888. doi:10.1016/S1053-8119(03)00336-7

31. Andersson JLR, Sotiropoulos SN. An integrated approach to correction for off-resonance effects and subject movement in diffusion MR imaging. NeuroImage. 2016;125:1063–1078. doi:10.1016/j.neuroimage.2015.10.019

32. Grabner G, Janke AL, Budge MM, Smith D, Pruessner J, Collins DL. Symmetric atlasing and model based segmentation: an application to the hippocampus in older adults. Med Image Comput Comput Assist Interv. 2006;9(Pt 2):58–66.

33. de Groot M, Vernooij MW, Klein S, et al. Improving alignment in Tract-based spatial statistics: Evaluation and optimization of image registration. NeuroImage. 2013;76:400–411. doi:10.1016/j.neuroimage.2013.03.015

34. Warrington S, Thompson E, Bastiani M, et al. Concurrent mapping of brain ontogeny and phylogeny within a common space: Standardized tractography and applications. Sci Adv. 2022;8(42):eabq2022. doi:10.1126/sciadv.abq2022

35. Warrington S, Bryant KL, Khrapitchev AA, et al. XTRACT - Standardised protocols for automated tractography in the human and macaque brain. Neuroimage. 2020;217:116923. doi:10.1016/j.neuroimage.2020.116923

36. Schmidt P, Gaser C, Arsic M, et al. An automated tool for detection of FLAIR-hyperintense white-matter lesions in Multiple Sclerosis. Neuroimage. 2012;59(4):3774–3783. doi:10.1016/j.neuroimage.2011.11.032

37. Pituch KA, Stevens JP. Applied Multivariate Statistics for the Social Sciences: Analyses with SAS and IBM’s SPSS, Sixth Edition. Routledge; 2015.

38. Youngstrom EA. A primer on receiver operating characteristic analysis and diagnostic efficiency statistics for pediatric psychology: we are ready to ROC. J Pediatr Psychol. 2014;39(2):204–221. doi:10.1093/jpepsy/jst062

39. Backner Y, Kuchling J, Massarwa S, et al. Anatomical Wiring and Functional Networking Changes in the Visual System Following Optic Neuritis. JAMA Neurology. 2018;75(3):287–295. doi:10.1001/jamaneurol.2017.3880

40. Kolbe SC, van der Walt A, Butzkueven H, Klistorner A, Egan GF, Kilpatrick TJ. Serial Diffusion Tensor Imaging of the Optic Radiations after Acute Optic Neuritis. J Ophthalmol. 2016;2016:2764538. doi:10.1155/2016/2764538

41. Kuchling J, Backner Y, Oertel FC, et al. Comparison of probabilistic tractography and tract-based spatial statistics for assessing optic radiation damage in patients with autoimmune inflammatory disorders of the central nervous system. Neuroimage Clin. 2018;19:538–550. doi:10.1016/j.nicl.2018.05.004

42. Tur C, Goodkin O, Altmann DR, et al. Longitudinal evidence for anterograde trans-synaptic degeneration after optic neuritis. Brain. 2016;139(Pt 3):816–828. doi:10.1093/brain/awv396

43. Gajamange S, Raffelt D, Dhollander T, et al. Fibre-specific white matter changes in multiple sclerosis patients with optic neuritis. Neuroimage Clin. 2018;17:60–68. doi:10.1016/j.nicl.2017.09.027

44. Kolbe S, Bajraszewski C, Chapman C, et al. Diffusion tensor imaging of the optic radiations after optic neuritis. Hum Brain Mapp. 2012;33(9):2047–2061. doi:10.1002/hbm.21343

45. Pawlitzki M, Horbrügger M, Loewe K, et al. MS optic neuritis-induced long-term structural changes within the visual pathway. Neurol Neuroimmunol Neuroinflamm. 2020;7(2):e665. doi:10.1212/NXI.0000000000000665

46. Raz N, Bick AS, Ben-Hur T, Levin N. Focal demyelinative damage and neighboring white matter integrity: an optic neuritis study. Mult Scler. 2015;21(5):562–571. doi:10.1177/1352458514551452

47. Horbruegger M, Loewe K, Kaufmann J, et al. Anatomically constrained tractography facilitates biologically plausible fiber reconstruction of the optic radiation in multiple sclerosis. Neuroimage Clin. 2019;22:101740. doi:10.1016/j.nicl.2019.101740

48. Ngamsombat C, Tian Q, Fan Q, et al. Axonal damage in the optic radiation assessed by white matter tract integrity metrics is associated with retinal thinning in multiple sclerosis. Neuroimage Clin. 2020;27:102293. doi:10.1016/j.nicl.2020.102293

49. Behrens TEJ, Berg HJ, Jbabdi S, Rushworth MFS, Woolrich MW. Probabilistic diffusion tractography with multiple fibre orientations: What can we gain? NeuroImage. 2007;34(1):144–155. doi:10.1016/j.neuroimage.2006.09.018

50. Dayan M, Munoz M, Jentschke S, et al. Optic radiation structure and anatomy in the normally developing brain determined using diffusion MRI and tractography. Brain Struct Funct. 2015;220(1):291–306. doi:10.1007/s00429-013-0655-y

51. Honnedevasthana Arun A, Connelly A, Smith RE, Calamante F. Characterisation of white matter asymmetries in the healthy human brain using diffusion MRI fixel-based analysis. NeuroImage. 2021;225:117505. doi:10.1016/j.neuroimage.2020.117505

52. Scheel M, Finke C, Oberwahrenbrock T, et al. Retinal nerve fibre layer thickness correlates with brain white matter damage in multiple sclerosis: a combined optical coherence tomography and diffusion tensor imaging study. Mult Scler. 2014;20(14):1904–1907. doi:10.1177/1352458514535128

53. Winklewski PJ, Sabisz A, Naumczyk P, Jodzio K, Szurowska E, Szarmach A. Understanding the Physiopathology Behind Axial and Radial Diffusivity Changes— What Do We Know? Front Neurol. 2018;9:92. doi:10.3389/fneur.2018.00092

