## Supplementary Material for "Diffusion-weighted Imaging And Retinal Oximetry Predict Functional Outcome After The First Episode Of Optic Neuritis"

### Supplementary Tables

**Supplementary Table 1. Summary of outcome measures, regressors and statistical tests**

| Number | Hypothesis | Outcome measures (dependent variables) | Explanatory variables | Confounders | Statistical Test |
|---|---|---|---|---|---|
| **1** | Group differences in DWI parameters | FA, MD, AD, RD[a], f1[a], f2 (LOR/ROR) at M0 | group, hemisphere | none | MANOVA |
| | | | | age, sex | |
| **2** | Change in DWI parameters over time | FA, MD, AD, RD[a], f1[a], f2 (LOR/ROR) at M0, M3, and M6 | time, hemisphere | none | MANOVA |
| | | | | affected side, time since onset | |
| | | | time, hemisphere, abnormal outcome | none | MANOVA |
| **3** | Correlation with clinical parameters and retinal imaging | BCVA, Pelli-Robson score, RNFL AS, VS, AVD of the AE at M0, M3, and M6 | FA, MD, AD, RD, f1, f2 (LOR/ROR) at M0, M3, and M6 | none | Spearman's rank correlation coefficient |
| **4** | Prediction of clinical parameters and retinal imaging at M6 | BCVA, Pelli-Robson score, RNFL AS, VS, AVD of the AE at M6 | FA, MD, AD, RD, f1, f2 (LOR/ROR) at M0 | none | Spearman's rank correlation coefficient |
| | | | **Auxiliary hypotheses** | | |
| **5** | Correlation between clinical parameters and retinal imaging | BCVA, Pelli-Robson score of the AE at M0, M3, and M6 | RNFL AS, VS, AVD of the AE at M0, M3, and M6 | none | Spearman's rank correlation coefficient |
| **6** | Prediction of clinical parameters at M6 | BCVA, Pelli-Robson score of the AE at M6 | RNFL AS, VS, AVD of the AE at M0 | none | Spearman's rank correlation coefficient |
| **7** | Prediction of retinal atrophy at M6 | RNFL of the AE at M6 | AS, VS, AVD of the AE at M0 | none | Spearman's rank correlation coefficient |
| **8** | Correlation with lesion load | FA, MD, AD, RD, f1, f2 (LOR/ROR) at M0, M3, and M6 | LL at M0, M3, and M6 | none | Spearman's rank correlation coefficient |

**Notes:** [a)]Variables excluded from the main analysis due to multicollinearity.

**Abbreviations:** AD – axonal diffusivity; AE – affected eye; AS – arteriolar (oxygen) saturation; AVD – arterio-venous difference; BCVA – Best-Corrected Visual Acuity; f1 – primary partial volume fraction; f2 – secondary partial volume fraction; FA – fractional anisotropy; LL – lesion load; LOR – left optic radiation; M0 – month 0 (baseline); M3 – month 3; M6 – month 6; MANOVA – multivariate analysis of variance; MD – mean diffusivity; RD – radial diffusivity; RNFL – retinal nerve fiber layer; ROR – right optic radiation; VS – venular (oxygen) saturation.

**Supplementary Table 2. Summary DWI measures**

| | | Patients with optic neuritis | | | | | | HCs | |
|---|---|---|---|---|---|---|---|---|---|
| | | M0[a] | | M3[b] | | M6[b] | | M0 | |
| ***n*** | | 24 | | 17 | | 17 | | 56 | |
| | | **Mean** | **SD** | **Mean** | **SD** | **Mean** | **SD** | **Mean** | **SD** |
| **FA** | **L** | 0.57 | 0.03 | 0.57 | 0.03 | 0.57 | 0.03 | 0.58 | 0.02 |
| | **R** | 0.56 | 0.03 | 0.56 | 0.03 | 0.56 | 0.03 | 0.57 | 0.02 |
| **MD [$\times 10^{-3}$ mm$^2$/s]** | **L** | 0.81 | 0.03 | 0.81 | 0.03 | 0.80 | 0.03 | 0.80 | 0.03 |
| | **R** | 0.79 | 0.03 | 0.79 | 0.02 | 0.79 | 0.03 | 0.78 | 0.03 |
| **AD [$\times 10^{-3}$ mm$^2$/s]** | **L** | 1.40 | 0.05 | 1.39 | 0.05 | 1.38 | 0.05 | 1.40 | 0.05 |
| | **R** | 1.34 | 0.05 | 1.34 | 0.04 | 1.34 | 0.04 | 1.35 | 0.05 |
| **RD [$\times 10^{-3}$ mm$^2$/s]** | **L** | 0.51 | 0.03 | 0.51 | 0.03 | 0.51 | 0.03 | 0.50 | 0.03 |
| | **R** | 0.51 | 0.03 | 0.51 | 0.03 | 0.51 | 0.03 | 0.50 | 0.03 |
| **f1** | **L** | 0.45 | 0.03 | 0.44 | 0.03 | 0.44 | 0.03 | 0.46 | 0.02 |
| | **R** | 0.43 | 0.03 | 0.43 | 0.02 | 0.44 | 0.02 | 0.44 | 0.02 |
| **f2** | **L** | 0.13 | 0.01 | 0.13 | 0.01 | 0.13 | 0.01 | 0.12 | 0.02 |
| | **R** | 0.15 | 0.02 | 0.15 | 0.01 | 0.14 | 0.01 | 0.14 | 0.02 |

**Notes:** [a)]all included subjects; [b)]PwON included in longitudinal multivariate analysis of variance.

**Abbreviations:** AD – axonal diffusivity; DWI – diffusion-weighted imaging; f1 – primary partial volume fraction; f2 – secondary partial volume fraction; FA – fractional anisotropy; HCs – healthy controls; L – left; MD – mean diffusivity; R – right; RD – radial diffusivity; SD – standard deviation.

**Supplementary Table 3. MANOVA and post-hoc ANOVA for group comparison at baseline (analysis with f1)**

| | MANOVA | | | | | | ANOVA | | | | |
|---|---|---|---|---|---|---|---|---|---|---|---|
| **Model** | **factor** | **Wilk's λ** | **df$^a$** | ***F*** | ***p*** | **Partial $\eta^2$** | **Variable** | **df$^a$** | ***F*** | ***p$^b$*** | **Partial $\eta^2$** |
| **Unadjusted 24 patients vs. 56 HCs** | group | 0.841 | 4, 75 | 3.554 | **0.010** | 0.159 | MD | 1, 78 | 0.639 | 0.427 | 0.008 |
| | | | | | | | AD | 1, 78 | 0.148 | 0.702 | 0.002 |
| | | | | | | | f1 | 1, 78 | 4.147 | 0.045 | 0.050 |
| | | | | | | | f2 | 1, 78 | 8.799 | **0.004** | 0.101 |
| | hemisphere | 0.325 | 4, 75 | 38.857 | **<0.001** | 0.675 | MD | 1, 78 | 49.453 | **<0.001** | 0.388 |
| | | | | | | | AD | 1, 78 | 138.484 | **<0.001** | 0.640 |
| | | | | | | | f1 | 1, 78 | 29.033 | **<0.001** | 0.271 |
| | | | | | | | f2 | 1, 78 | 81.230 | **<0.001** | 0.510 |
| | group*hemisphere | 0.949 | 4, 75 | 1.001 | 0.412 | 0.051 | | | | | |
| **Adjusted 24 patients vs. 56 HCs** | group | 0.904 | 4, 72 | 1.920 | 0.116 | 0.096 | | | | | |
| | hemisphere | 0.917 | 4, 72 | 1.632 | 0.175 | 0.083 | | | | | |
| | age | 0.934 | 4, 72 | 1.266 | 0.291 | 0.066 | | | | | |
| | sex | 0.790 | 4, 72 | 4.787 | **0.002** | 0.210 | MD | 1, 75 | 0.415 | 0.521 | 0.006 |
| | | | | | | | AD | 1, 75 | 4.185 | 0.044 | 0.053 |
| | | | | | | | f1 | 1, 75 | 2.688 | 0.105 | 0.035 |
| | | | | | | | f2 | 1, 75 | 16.643 | **<0.001** | 0.182 |
| | group*sex | 0.965 | 4, 72 | 0.654 | 0.626 | 0.035 | | | | | |
| | group*hemisphere | 0.972 | 4, 72 | 0.514 | 0.725 | 0.028 | | | | | |
| | hemisphere*age | 0.976 | 4, 72 | 0.449 | 0.772 | 0.024 | | | | | |
| | hemisphere*sex | 0.992 | 4, 72 | 0.147 | 0.964 | 0.008 | | | | | |
| | group*hemisphere*sex | 0.992 | 4, 72 | 0.140 | 0.967 | 0.008 | | | | | |
| **Unadjusted 24 patients vs. 24 HCs** | group | 0.759 | 4, 43 | 3.412 | **0.016** | 0.241 | MD | 1, 46 | 0.533 | 0.469 | 0.011 |
| | | | | | | | AD | 1, 46 | 1.266 | 0.266 | 0.027 |
| | | | | | | | f1 | 1, 46 | 6.437 | 0.015 | 0.123 |
| | | | | | | | f2 | 1, 46 | 7.795 | **0.008** | 0.145 |
| | hemisphere | 0.305 | 4, 43 | 24.542 | **<0.001** | 0.695 | MD | 1, 46 | 41.767 | **<0.001** | 0.476 |
| | | | | | | | AD | 1, 46 | 93.242 | **<0.001** | 0.670 |
| | | | | | | | f1 | 1, 46 | 20.792 | **<0.001** | 0.311 |
| | | | | | | | f2 | 1, 46 | 48.213 | **<0.001** | 0.512 |
| | group*hemisphere | 0.934 | 4, 43 | 0.757 | 0.559 | 0.066 | | | | | |

**Notes:** [a]Order: hypothesis df, error df; [b]Bonferroni-corrected significance level $p < 0.0125$ for $n = 4$ marked in **bold**.

**Abbreviations:** AD – axonal diffusivity; ANOVA – analysis of variance; df – degrees of freedom; f1 – primary partial volume fraction; f2 – secondary partial volume fraction; FA – fractional anisotropy; HCs – healthy controls; MANOVA – multivariate ANOVA; MD – mean diffusivity.

**Supplementary Table 4. MANOVA and post-hoc ANOVA for longitudinal assessment in patients, irrespective of outcome (analysis with f1)**

| | MANOVA | | | | | | ANOVA | | | | |
|---|---|---|---|---|---|---|---|---|---|---|---|
| **Model** | **factor** | **Wilk's λ** | **df[a]** | ***F*** | ***p*** | **Partial $\eta^2$** | **Variable** | **df[a]** | ***F*** | **$p^b$** | **Partial $\eta^2$** |
| Unadjusted | time | 0.519 | 8, 9 | 1.041 | 0.472 | 0.481 | | | | | |
| | hemisphere | 0.209 | 4, 13 | 12.279 | **<0.001** | 0.791 | MD | 1, 16 | 15.308 | **0.001** | 0.489 |
| | | | | | | | AD | 1, 16 | 38.641 | **<0.001** | 0.707 |
| | | | | | | | f1 | 1, 16 | 3.977 | 0.063 | 0.199 |
| | | | | | | | f2 | 1, 16 | 38.017 | **<0.001** | 0.704 |
| | time*hemisphere | 0.354 | 8, 9 | 2.056 | 0.152 | 0.646 | | | | | |
| Adjusted | time | 0.190 | 8, 7 | 3.729 | **0.050** | 0.810 | MD | 2, 28 | 2.555 | 0.096 | 0.154 |
| | | | | | | | AD | 2, 28 | 4.294 | 0.024 | 0.235 |
| | | | | | | | f1 | 2, 28 | 1.636 | 0.213 | 0.105 |
| | | | | | | | f2 | 2, 28 | 0.580 | 0.566 | 0.040 |
| | hemisphere | 0.445 | 4, 11 | 3.424 | **0.047** | 0.555 | MD | 1, 14 | 13.039 | **0.003** | 0.482 |
| | | | | | | | AD | 1, 14 | 12.035 | **0.004** | 0.462 |
| | | | | | | | f1 | 1, 14 | 0.005 | 0.943 | 0.000 |
| | | | | | | | f2 | 1, 14 | 6.136 | 0.027 | 0.305 |
| | affected side | 0.604 | 4, 11 | 1.804 | 0.198 | 0.396 | | | | | |
| | time since onset | 0.754 | 4, 11 | 0.897 | 0.498 | 0.246 | | | | | |
| | time*hemisphere | 0.412 | 8, 7 | 1.249 | 0.391 | 0.588 | | | | | |
| | time*affected side | 0.189 | 8, 7 | 3.763 | **0.049** | 0.811b | MD | 2, 28 | 1.438 | 0.254 | 0.093 |
| | | | | | | | AD | 2, 28 | 2.448 | 0.105 | 0.149 |
| | | | | | | | f1 | 2, 28 | 1.435 | 0.255 | 0.093 |
| | | | | | | | f2 | 2, 28 | 0.046 | 0.955 | 0.003 |
| | time* time since onset | 0.213 | 8, 7 | 3.231 | 0.070 | 0.787 | | | | | |
| | hemisphere* affected side | 0.882 | 4, 11 | 0.367 | 0.827 | 0.118 | | | | | |
| | hemisphere* time since onset | 0.679 | 4, 11 | 1.303 | 0.328 | 0.321 | | | | | |
| | time*hemisphere* affected side | 0.347 | 8, 7 | 1.645 | 0.263 | 0.653 | | | | | |
| | time*hemisphere* time since onset | 0.442 | 8, 7 | 1.103 | 0.455 | 0.558 | | | | | |

**Notes:** a)Order: hypothesis df, error df; b)Bonferroni-corrected significance level $p < 0.0125$ for $n = 4$ marked in **bold**.

**Abbreviations:** AD – axonal diffusivity; ANOVA – analysis of variance; df – degrees of freedom; f2 – secondary partial volume fraction; FA – fractional anisotropy; MANOVA – multivariate ANOVA; MD – mean diffusivity.

**Supplementary Table 5. MANOVA and ANOVA for longitudinal assessment in patients, stratified by outcome (analysis with f1)**

| | | MANOVA | | | | | ANOVA | | | | |
|---|---|---|---|---|---|---|---|---|---|---|---|
| **Model** | **factor** | **Wilk's λ** | **df[a]** | ***F*** | ***p*** | **Partial $\eta^2$** | **Variable** | **df[a]** | ***F*** | **$p^b$** | **Partial $\eta^2$** |
| **Stratified by outcome** | abnormal outcome | 0.342 | 4, 12 | 5.782 | **0.008** | 0.658 | MD | 1, 15 | 0.424 | 0.525 | 0.027 |
| | | | | | | | AD | 1, 15 | 3.037 | 0.102 | 0.168 |
| | | | | | | | f1 | 1, 15 | 4.650 | 0.048 | 0.237 |
| | | | | | | | f2 | 1, 15 | 1.566 | 0.230 | 0.095 |
| | time | 0.518 | 8, 8 | 0.929 | 0.540 | 0.482 | | | | | |
| | hemisphere | 0.251 | 4, 12 | 8.935 | **0.001** | 0.749 | MD | 1, 15 | 11.436 | **0.004** | 0.433 |
| | | | | | | | AD | 1, 15 | 28.808 | **<0.001** | 0.658 |
| | | | | | | | f1 | 1, 15 | 2.681 | 0.122 | 0.152 |
| | | | | | | | f2 | 1, 15 | 27.090 | **<0.001** | 0.644 |
| | abnormal outcome* time | 0.469 | 8, 8 | 1.134 | 0.432 | 0.531 | | | | | |
| | abnormal outcome* hemisphere | 0.843 | 4, 12 | 0.561 | 0.696 | 0.157 | | | | | |
| | time*hemisphere | 0.157 | 8, 8 | 5.388 | **0.014** | 0.843 | MD | 2, 30 | 1.041 | 0.365 | 0.065 |
| | | | | | | | AD | 2, 30 | 1.270 | 0.295 | 0.078 |
| | | | | | | | f1[c] | 2, 30 | 2.818 | 0.095 | 0.158 |
| | | | | | | | f2 | 2, 30 | 0.855 | 0.435 | 0.054 |
| | abnormal outcome* time*hemisphere | 0.235 | 8, 8 | 3.259 | 0.057 | 0.765 | | | | | |

**Notes:** [a)]Order: hypothesis df, error df; [b)]Bonferroni-corrected significance level $p < 0.0125$ for $n = 4$ marked in **bold**; [c)]Greenhouse-Geisser corrected univariate statistics.

**Abbreviations:** AD – axonal diffusivity; ANOVA – analysis of variance; df – degrees of freedom; f2 – secondary partial volume fraction; FA – fractional anisotropy; MANOVA – multivariate ANOVA; MD – mean diffusivity.

**Supplementary Table 6. Correlations between DWI and ophthalmological parameters at each time point**

| | | | BCVA | | Pelli-Robson | | RNFL | | AS | | VS | | AVD | |
|---|---|---|---|---|---|---|---|---|---|---|---|---|---|---|
| ***n*** | | | 23 | | 15 | | 23 | | 22 | | 22 | | 22 | |
| | | | $\varrho^a$ | $p^b$ | $\varrho^a$ | $p^b$ | $\varrho^a$ | $p^b$ | $\varrho^a$ | $p^b$ | $\varrho^a$ | $p^b$ | $\varrho^a$ | $p^b$ |
| **M0** | **FA** | **L** | 0.074 | 0.736 | 0.059 | 0.833 | *0.442* | *0.035* | 0.174 | 0.440 | 0.219 | 0.328 | 0.089 | 0.694 |
| | | **R** | 0.180 | 0.411 | 0.097 | 0.730 | 0.153 | 0.486 | 0.248 | 0.265 | 0.314 | 0.154 | −0.057 | 0.802 |
| | **MD** | **L** | 0.178 | 0.416 | 0.095 | 0.735 | −0.320 | 0.137 | −0.095 | 0.675 | −0.203 | 0.366 | −0.005 | 0.982 |
| | | **R** | 0.135 | 0.539 | −0.087 | 0.758 | −0.213 | 0.329 | −0.010 | 0.964 | 0.027 | 0.906 | −0.253 | 0.257 |
| | **AD** | **L** | 0.202 | 0.355 | 0.205 | 0.464 | −0.016 | 0.943 | 0.100 | 0.659 | −0.058 | 0.798 | 0.052 | 0.820 |
| | | **R** | 0.286 | 0.186 | 0.088 | 0.754 | −0.154 | 0.484 | 0.158 | 0.484 | 0.095 | 0.675 | −0.252 | 0.259 |
| | **RD** | **L** | 0.057 | 0.797 | −0.013 | 0.964 | *−0.423* | *0.044* | −0.073 | 0.746 | −0.235 | 0.292 | −0.029 | 0.899 |
| | | **R** | −0.002 | 0.994 | −0.136 | 0.628 | −0.294 | 0.173 | −0.144 | 0.523 | −0.125 | 0.579 | −0.156 | 0.488 |
| | **f1** | **L** | 0.002 | 0.993 | −0.002 | 0.995 | *0.446* | *0.033* | 0.178 | 0.429 | 0.208 | 0.353 | 0.099 | 0.663 |
| | | **R** | 0.093 | 0.674 | 0.031 | 0.914 | 0.182 | 0.405 | 0.245 | 0.272 | 0.228 | 0.308 | 0.071 | 0.754 |
| | **f2** | **L** | −0.105 | 0.633 | −0.142 | 0.613 | 0.125 | 0.571 | −0.162 | 0.473 | −0.229 | 0.304 | 0.216 | 0.335 |
| | | **R** | 0.080 | 0.717 | 0.079 | 0.779 | 0.375 | 0.078 | −0.226 | 0.313 | 0.055 | 0.808 | −0.021 | 0.926 |
| ***n*** | | | 20 | | 15 | | 20 | | 19 | | 19 | | 19 | |
| **M3** | **FA** | **L** | 0.141 | 0.553 | 0.328 | 0.232 | 0.089 | 0.709 | 0.216 | 0.375 | 0.410 | 0.081 | −0.304 | 0.205 |
| | | **R** | 0.123 | 0.605 | *0.603* | *0.017* | 0.165 | 0.487 | 0.006 | 0.980 | −0.019 | 0.940 | −0.021 | 0.932 |
| | **MD** | **L** | 0.075 | 0.753 | 0.069 | 0.808 | 0.151 | 0.526 | 0.110 | 0.653 | −0.171 | 0.484 | 0.241 | 0.320 |
| | | **R** | −0.035 | 0.882 | −0.300 | 0.276 | 0.219 | 0.353 | −0.161 | 0.511 | −0.220 | 0.366 | 0.187 | 0.442 |
| | **AD** | **L** | 0.228 | 0.333 | 0.223 | 0.425 | 0.293 | 0.211 | 0.115 | 0.640 | 0.266 | 0.271 | −0.156 | 0.524 |
| | | **R** | 0.022 | 0.926 | 0.204 | 0.466 | 0.251 | 0.286 | −0.239 | 0.325 | −0.187 | 0.443 | 0.074 | 0.764 |
| | **RD** | **L** | −0.034a | 0.888 | −0.115 | 0.683 | −0.053 | 0.825 | −0.093 | 0.704 | −0.269 | 0.265 | 0.230 | 0.344 |
| | | **R** | 0.004 | 0.985 | *−0.607* | *0.017* | 0.034 | 0.887 | −0.116 | 0.637 | −0.169 | 0.490 | 0.152 | 0.534 |
| | **f1** | **L** | 0.139 | 0.558 | 0.378 | 0.164 | 0.038 | 0.875 | 0.214 | 0.379 | 0.360 | 0.130 | −0.279 | 0.248 |
| | | **R** | 0.039 | 0.870 | *0.623* | *0.013* | 0.058 | 0.808 | 0.006 | 0.980 | 0.049 | 0.841 | −0.074 | 0.764 |
| | **f2** | **L** | −0.277 | 0.238 | −0.043 | 0.880 | 0.221 | 0.349 | −0.185 | 0.447 | −0.301 | 0.210 | 0.199 | 0.414 |
| | | **R** | 0.233 | 0.322 | 0.189 | 0.499 | 0.153 | 0.519 | −0.037 | 0.880 | −0.057 | 0.818 | −0.027 | 0.912 |
| ***n*** | | | 18 | | 15 | | 18 | | 18 | | 18 | | 18 | |
| **M6** | **FA** | **L** | 0.385 | 0.114 | 0.479 | 0.071 | −0.141 | 0.576 | 0.172 | 0.495 | 0.015 | 0.954 | 0.162 | 0.520 |
| | | **R** | 0.271 | 0.277 | 0.485 | 0.067 | −0.137 | 0.587 | 0.297 | 0.232 | −0.079 | 0.756 | 0.175 | 0.488 |
| | **MD** | **L** | −0.182 | 0.471 | −0.065 | 0.818 | −0.174 | 0.491 | −0.058 | 0.818 | −0.355 | 0.149 | 0.208 | 0.408 |
| | | **R** | −0.184 | 0.464 | −0.211 | 0.451 | −0.035 | 0.890 | −0.304 | 0.220 | −0.389 | 0.110 | 0.188 | 0.455 |
| | **AD** | **L** | 0.405 | 0.095 | 0.479 | 0.071 | −0.022 | 0.932 | 0.058 | 0.820 | −0.235 | 0.347 | 0.271 | 0.277 |
| | | **R** | 0.119 | 0.638 | 0.249 | 0.371 | −0.121 | 0.633 | 0.023 | 0.926 | *−0.553* | *0.017* | *0.515* | *0.029* |
| | **RD** | **L** | −0.150 | 0.554 | −0.306 | 0.217 | 0.071 | 0.778 | 0.086 | 0.735 | −0.299 | 0.229 | −0.375 | 0.168 |
| | | **R** | −0.357 | 0.146 | −0.187 | 0.458 | −0.004 | 0.987 | 0.173 | 0.491 | −0.139 | 0.583 | −0.302 | 0.273 |
| | **f1** | **L** | 0.263 | 0.291 | 0.036 | 0.886 | 0.213 | 0.397 | −0.119 | 0.639 | 0.424 | 0.080 | 0.477 | 0.072 |
| | | **R** | 0.249 | 0.319 | 0.127 | 0.616 | 0.005 | 0.984 | −0.199 | 0.428 | 0.201 | 0.423 | *0.564* | *0.028* |
| | **f2** | **L** | −0.125 | 0.622 | 0.160 | 0.570 | −0.076 | 0.763 | −0.434 | 0.072 | −0.119 | 0.637 | −0.151 | 0.550 |
| | | **R** | 0.292 | 0.240 | −0.187 | 0.504 | *0.570* | *0.014* | −0.159 | 0.528 | 0.181 | 0.473 | −0.458 | 0.056 |

**Notes:** [a)]Spearman rank correlation coefficient; [b)]Significant correlations at Bonferroni-Holm-corrected significance level p < 0.0083 ($n = 6$) marked in **bold**, uncorrected significance ($p < 0.05$) marked in *italics*.

**Abbreviations:** AD – axonal diffusivity; AS – arteriolar (oxygen) saturation; AVD – arterio-venous difference; BCVA – Best-Corrected Visual Acuity; DWI – diffusion-weighted imaging; f2 – secondary partial volume fraction; FA – fractional anisotropy; L – left; M0 – visit at month 0 (baseline); M3 – visit at month 3; M6 – visit at month 6; MD – mean diffusivity; R – right; RNFL – retinal nerve fiber layer; VS – venular (oxygen) saturation.

**Supplementary Table 7. Correlations among retinal parameters and visual function**

| | | BCVA | | | Pelli-Robson | | | RNFL | | |
|---|---|---|---|---|---|---|---|---|---|---|
| | | *ϱ*[a] | *p* | *n* | *ϱ*[a] | *p* | *n* | *ϱ*[a] | *p* | *n* |
| | | | | | **M0** | | | | | |
| **M0** | **RNFL** | 0.039 | 0.861 | 23 | −0.010 | 0.972 | 15 | | | |
| | **AS** | *−0.390* | *0.073* | 22 | *−0.490* | *0.063* | 15 | | | |
| | **VS** | 0.223 | 0.318 | 22 | −0.229 | 0.412 | 15 | | | |
| | **AVD** | **−0.604** | **0.003** | 22 | −0.322 | 0.242 | 15 | | | |
| | | | | | **M3** | | | | | |
| **M3** | **RNFL** | 0.260 | 0.269 | 20 | −0.098 | 0.729 | 15 | | | |
| | **AS** | 0.052 | 0.833 | 19 | −0.158 | 0.589 | 14 | | | |
| | **VS** | −0.304 | 0.205 | 19 | 0.039 | 0.894 | 14 | | | |
| | **AVD** | 0.312 | 0.194 | 19 | −0.156 | 0.594 | 14 | | | |
| | | | | | **M6** | | | | | |
| **M6** | **RNFL** | **0.495** | **0.031** | 19 | 0.032 | 0.906 | 16 | | | |
| | **AS** | −0.006 | 0.980 | 19 | −0.217 | 0.420 | 16 | | | |
| | **VS** | −0.314 | 0.190 | 19 | −0.372 | 0.156 | 16 | | | |
| | **AVD** | 0.190 | 0.437 | 19 | 0.166 | 0.538 | 16 | | | |
| | | | | | **M6 (prediction)** | | | | | |
| **M0** | **RNFL** | 0.273 | 0.257 | 19 | 0.041 | 0.881 | 16 | 0.047 | 0.848 | 19 |
| | **AS** | 0.179 | 0.477 | 18 | 0.239 | 0.391 | 15 | −0.074 | 0.769 | 18 |
| | **VS** | **0.609** | **0.007** | 18 | 0.155 | 0.582 | 15 | **0.571** | **0.013** | 18 |
| | **AVD** | *−0.410* | *0.091* | 18 | 0.109 | 0.698 | 15 | **−0.747** | **<0.001** | 18 |

**Notes:** [a)]Spearman rank correlation coefficient; [b)]Uncorrected statistics. Significant correlations ($p < 0.05$) marked in bold, trends ($p < 0.10$) marked in italics.

**Abbreviations:** AD – axonal diffusivity; AS – arteriolar (oxygen) saturation; AVD – arterio-venous difference; BCVA – Best-Corrected Visual Acuity; DWI – diffusion-weighted imaging; f2 – secondary partial volume fraction; FA – fractional anisotropy; L – left; M0 – visit at month 0 (baseline); M3 – visit at month 3; M6 – visit at month 6; MD – mean diffusivity; N/A – not applicable; PR – Pelli-Robson score; R – right; RNFL – retinal nerve fiber layer; VS – venular (oxygen) saturation.

**Supplementary Table 8. Correlations between DWI parameters and LL**

| | | | Whole-brain LL | | OR lesion volume fraction | |
|---|---|---|---|---|---|---|
| | | | **M0 ($n$ = 24)** | | | |
| | | | $\varrho^a$ | $p^b$ | $\varrho^a$ | $p^b$ |
| **M0** | **FA** | **L** | −0.086 | 0.689 | −0.155 | 0.471 |
| | | **R** | −0.113 | 0.600 | −0.031 | 0.886 |
| | **MD** | **L** | 0.241 | 0.258 | 0.235 | 0.270 |
| | | **R** | 0.120 | 0.576 | 0.000 | 0.998 |
| | **AD** | **L** | 0.183 | 0.392 | 0.189 | 0.378 |
| | | **R** | 0.067 | 0.756 | −0.130 | 0.546 |
| | **RD** | **L** | 0.240 | 0.259 | 0.212 | 0.319 |
| | | **R** | 0.122 | 0.569 | 0.045 | 0.835 |
| | **f1** | **L** | −0.046 | 0.832 | −0.163 | 0.446 |
| | | **R** | −0.154 | 0.471 | −0.015 | 0.943 |
| | **f2** | **L** | −0.200 | 0.349 | −0.189 | 0.377 |
| | | **R** | −0.037 | 0.864 | −0.004 | 0.987 |
| | | | **M3 ($n$ = 21)** | | | |
| **M3** | **FA** | **L** | −0.008 | 0.973 | −0.354 | 0.115 |
| | | **R** | −0.044 | 0.849 | −0.258 | 0.258 |
| | **MD** | **L** | 0.357 | 0.113 | 0.378 | 0.091 |
| | | **R** | 0.060 | 0.797 | 0.000 | 1.000 |
| | **AD** | **L** | 0.248 | 0.279 | −0.070 | 0.762 |
| | | **R** | 0.034 | 0.884 | −0.222 | 0.334 |
| | **RD** | **L** | 0.240 | 0.294 | 0.382 | 0.087 |
| | | **R** | 0.048 | 0.836 | 0.259 | 0.258 |
| | **f1** | **L** | −0.030 | 0.898 | −0.354 | 0.115 |
| | | **R** | −0.055 | 0.814 | −0.295 | 0.194 |
| | **f2** | **L** | −0.269 | 0.239 | −0.115 | 0.621 |
| | | **R** | 0.081 | 0.729 | 0.369 | 0.099 |
| | | | **M6 ($n$ = 18)** | | | |
| **M6** | **FA** | **L** | 0.018 | 0.945 | −0.189 | 0.453 |
| | | **R** | 0.020 | 0.938 | N/A[c] | |
| | **MD** | **L** | 0.447 | 0.063 | 0.272 | 0.276 |
| | | **R** | 0.160 | 0.526 | N/A[c] | |
| | **AD** | **L** | 0.277 | 0.266 | −0.017 | 0.947 |
| | | **R** | 0.127 | 0.616 | N/A[c] | |
| | **RD** | **L** | 0.159 | 0.528 | 0.257 | 0.303 |
| | | **R** | 0.061 | 0.810 | N/A[c] | |
| | **f1** | **L** | −0.112 | 0.657 | −0.193 | 0.444 |
| | | **R** | 0.042 | 0.868 | N/A[c] | |
| | **f2** | **L** | 0.290 | 0.243 | −0.266 | 0.285 |
| | | **R** | 0.053 | 0.836 | N/A[c] | |

**Notes:** [a)]Spearman rank correlation coefficient; [b)]Significant correlations uncorrected significance level $p < 0.05$ marked in **bold**; [c)]Lesion volume fraction for the right OR was 0 in all patients.

**Abbreviations:** AD – axonal diffusivity; DWI – diffusion-weighted imaging; f2 – secondary partial volume fraction; FA – fractional anisotropy; L – left; LL – lesion load; M0 – visit at month 0 (baseline); M3 – visit at month 3; M6 – visit at month 6; OR – optic radiation; R – right.
